# Mental health in the short- and long-term adaptation processes of university students during the COVID-19 pandemic: A systematic review and meta-analysis

**DOI:** 10.1101/2025.03.14.25323978

**Authors:** María Paola Jiménez-Villamizar, Laura Comendador Vázquez, Juan P Sanabria-Mazo, Corel Mateo, Josep María Losilla, Anna Muro, Antoni Sanz

**Author notes:** **Corresponding author** Carrer de la Fortuna, s/n, 08193 – Campus de Bellaterra, Cerdanyola del Vallès, Spain, /.

## Abstract

**Introduction:** During Covid-19, high prevalence of distress was reported among students, suggesting that they may be at higher risk than the general population of developing psychological disorders in confinement situations. **Methods:** We conducted a systematic search of four databases (PsycINFO, PubMed, SCOPUS, and Web of Science) for articles published from January 2020 to May 2022. Risk of bias was assessed using the Joanna Briggs Institute (JBI) checklist. Random effects meta-analyses of the reported proportions of college students with clinically significant symptoms of anxiety, depression and stress were carried out, and between-studies heterogeneity was also analysed. **Results:** 73 studies (N=209.761) were included for meta-analysis. The estimated proportion of college students with clinically significant short-term symptoms was 34% for anxiety (95% CI [29%,39%]; *I^2^*=99.75%), 38% for depression (95% CI [33%,44%; *I^2^* = 99.71%), and 54% for stress (95% CI [46%,62%]; *I^2^* = 99.57%). The estimated proportion of college students with clinically significant long-term symptoms was 37% for anxiety (95% CI [32%,42%]; *I^2^* = 97.92%), 31% for depression (95% CI [23%,41%]; *I^2^* = 99.49%) and 41% for stress (95% CI [25%,59%]; *I^2^* = 99.29%) were found. Several methodological and sociodemographic moderators accounted for heterogeneity in the observed prevalences. **Limitations:** The heterogeneity of study findings suggest that the results should be interpreted with caution. **Conclusion:** The current evidence shows that approximately one-third of college students experienced distress, further where we can infer that there was no evidence of a worsening in mental health derived from a cumulative effect during the pandemic.

PROSPERO: CRD420222233036.

## 1. Introduction

Pandemic situations, especially the exponential increase in cases of infection and death, generate fear of disease and uncertainty. This context meant that the vast majority of the population had to face stressful situations not previously experienced. During the COVID-19 public health emergency declared by WHO in 2020, the most of countries abruptly implemented a range of measures to mitigate the spread of the virus, as social distancing, isolation, and mobility restrictions, with complete closures and quarantines, which have forced citizens to stay at home for months (1,2). One of the sectors most affected by these sudden changes has been the educational sector, forcing institutions, universities, and schools to close their doors and rearrange their learning processes with online methods that deeply affected the daily routines of students and staff (3). The mental and emotional well-being of the younger student population may be altered in those countries that have been most affected by the pandemic. It is well known that prior to the COVID-19 crisis there were already economic and occupational concerns that were reflected in increased distress and mental health problems among university students (4), and the pandemic could have acted as a life event with the potentiality to increase the prevalence of mood disorders, aggravating a problem of progressive deterioration in the mental health of the younger generations, which had started previously (5).

Since 2020, many studies have been conducted in different countries on the psychological impact of the pandemic, most of them pointing to the enhanced vulnerability of younger populations (5,6). Among them, university students were affected by additional risk factors with the potential to increase the incidence of mental health problems, such as the difficulty and uncertainty related to the development of academic activities, the demands due to forced migration to online teaching and the new pedagogical dynamics (7). Literature reviews in university students report anxiety prevalence of 36%, depression of 39% (8), and stress between 28% and 70% (9). This purportedly confirms that the pandemic supposed the increase of negative emotions and psychological problems in this population group. The results of the present study seek to update the situation of the impact on mental health in university students in the face of COVID-19 collecting large-scale evidence on the distress of university students in two moments in front of an acute response that will be associated with the first wave of pandemic, while the second is associated with a time of evaluation of approximately second semester after the first wave (10). Distress is understood as chronic and prolonged emotional manifestations of discomfort, affecting adaptive capacity, and predisposing to the appearance of possible mental health problems (11). One of the studies found that less than half of the students in their sample were able to cope with this level of stress, while a significant proportion of students also reported having depressive thoughts (12), proposed reasons for influencing depressive symptoms include loneliness caused by social isolation and estrangement, power, financial uncertainty as major life reinforcers. Therefore, it is hypothesized that COVID-19 would have adverse effects on the mental health of students, the prevalence of these mental health problems will be higher in the acute phase than in the chronic phase would be affected by nations, gender and data collection period. Due to the heterogeneity in the results on changes in distress throughout the pandemic, systematization of the available literature seems to be necessary to understand the impact of COVID-19 on the mental health of college students. This systematic meta-analysis aimed to identify the prevalence of anxiety, depression, and stress in the short- and long-term coping processes of college students from a longitudinal perspective during the COVID-19 pandemic.

Review studies are needed to better describe and understand the psychological experience of university students and to facilitate health and educational policies to help this community cope with COVID-19 disruptions to their mental health and academic trajectory. Therefore, it is essential to study and review the current experience of students worldwide, the risk and protective factors, and the additional mental health costs of pandemic blockages, not only for the promotion of student mental health, but also for the design of systemic prevention and intervention programs in academic institutions (13,14).

### Current Study

The severity of distress symptoms such as depression and anxiety, as well as stress, has been reported in many studies on the psychological effects of the pandemic in university students, although they vary in the prevalence of intensity, being this group a priority focus of study due to the prevalences reported before and during the pandemic. No meta-analyses have been reported that inquire about the short-term and long-term evolution of mental health in college students during the COVID-19 pandemic. To address this gap, the aim of the present meta-analysis was to describe the short-term and long-term prevalence of depressive symptomatology, anxiety manifestations, and stress in college students during the COVID-19 pandemic.

The first objective was to identify and quantify indicators of distress according to the short-term adaptation process spanning from January 2020 to June 2020 (period 1) and long-term spanning from July 2020 to May 2022 (period 2). It should be noted that not all studies included in the current meta-analysis measured aspects of distress, therefore, all selected studies included (a) results of prevalences of depression, anxiety, or stress (b) date of data collection. As a secondary objective, moderation analyses were also applied to explore the effects of pandemic indicators (i.e., wave, confinement, severity), geographic location, and assessment instruments used. Such analyses were used to investigate sources of heterogeneity, which cannot be extracted from the pooled treatment effect estimate. Overall, the present study sought to assess the effects of short- and long-term prevalence of depressive symptomatology, manifestations of anxiety and stress in college students during the COVID-19 pandemic to establish recommendations for decision making on the implementation of effective programs and measures to promote a healthier college environment conducive to students’ intellectual and emotional growth.

## 2. Methods

### 2.1. Protocol and registration

The protocol was prepared in accordance with the Preferred Reporting Items for Protocols for Systematic Review and Meta-Analysis (PRISMA-P) (15,16). A version of the protocol was registered in the Prospective International Registry of Systematic Reviews (PROSPERO), with identification number CRD42022330361. This study was based on data collected from publicly available databases and did not require ethical approval from our institutional review boards.

### 2.2. Document search and selection

The literature search strategy was conducted in four electronic databases: PubMed (US National Library of Medicine), through the National Center for Biotechnology Information (NCBI), PsycInfo (American Psychological Association), through ProQuest, Scopus (Elsevier) and Web of Science Core Collection (Thomson Reuters) until June 26, 2022.

Empirical cross-sectional or longitudinal studies reporting the prevalence of depression-anxiety-stress in college students and/or examining through quantitative measures their associations with risk and protective factors were included in the review. Qualitative studies, individual case studies or case series, psychometric studies focused on instrument development or validation, opinion reports, and systematic reviews were excluded. For this purpose, it was essential to carry out a temporal delimitation between January 2020 and May 2022, and only articles in English and Spanish were reviewed. The search strategy was performed using the following PubMed search formula, modified according to the specific syntax of each of the bibliographic databases consulted (See Table S1).

In addition, we searched for literature included in the references, which was related to the topic and included in systematic reviews to complement, obtain relevant literature and ensure the retrieval ratio.

### 2.3. Eligibility criteria

Cross-sectional or longitudinal empirical studies that reported prevalence or mean mental health scores in college students were included in the review. Qualitative studies, single case studies or case series, psychometric studies focused on instrument validation, opinion reports, and systematic reviews were excluded. The PICOS-Population, Intervention, Comparison, Comparison, Outcome, or Outcome approach was followed to select eligibility criteria (17).

Inclusion criteria:

*Studies that report the prevalence of depression, anxiety or stress determined using a validated scale. The mean severity score of symptoms of depression, anxiety, or stress and its standard deviation among college students.

*Data collected during COVID-19, defined by the date of data collection taking into account two periods, one of short-term adaptation (data collected in the first waves of COVID-19 between January and June 2020), and another long-term (data collected from July 2020 to April 2022), taking into account the area or country where the study was conducted, this allowed to identify the temporal behaviour of the different waves of COVID (first, second or third wave). It is important to clarify that the pandemic wave consists of the growth of the number of sick people and ends with their decrease.

Exclusion criteria:

*Studies where data were collected prior to the onset of the COVID-19 pandemic were excluded.

*Studies where the population is from schools, primary and secondary educational institutions, or technicians.

### 2.4. Data management and study selection

In the first stage, the results of the review of all the databases were exported to Mendeley, where the software automatically eliminated duplicate articles in the databases, and then the first reviewer (MPJ) performed a manual review confirming the information. The second phase was performed through Rayyan (18) with a first screening from the title and abstract of the articles by two independent investigators (MPJ and LC). The initial inter-rater reliability Cohen’s kappa (19) was k = .92. In the third phase, the reviewers exported the selected references in Rayyan QCRI to a format developed by the team and proceeded to review the full texts to verify study eligibility. Finally, selected full text data, risk of bias (RoB) and methodological quality of included studies were extracted by the reviewers and key information was entered into a standardized database.

### 2.5. Data extraction and coding

Data extraction was performed independently by two authors (MPJ and LC), using a standard extraction form developed in a Microsoft Excel spreadsheet. Discrepancies were resolved by consensus and the participation of a third reviewer was not required.

Data were extracted from the following categories: (a) article identification data title (title, author, and year of publication); (b) sample characteristics (date of data collection, sample size, geographic location); (c) study measures anxiety, depression and/or stress; Methodological (instruments, reliability, mode of administration) and outcomes (mean, SD, prevalence, moderator variables).

For the analysis of the systematic review, we considered: (a) study identification data; (b) pandemic effects (COVID wave, isolation, pandemic severity); (c) sample characteristics (date of data collection, sample size, geographic location, percentage of women); (d) measures of anxiety, depression, and stress studies by period (number of subjects affected).

### 2.6. Risk of bias assessment

Risk of bias assessment was performed independently by two reviewers (MPJ and LC), using the Joanna Briggs Institute (JBI) Critical Appraisal Checklist for Studies Reporting Prevalence Data (20). Discrepancies were resolved by consensus and the involvement of a third reviewer was not necessary.

The JBI checklist for prevalence studies evaluates nine domains, in this case an adjustment was made where seven domains were evaluated: (1) adequacy of the sample frame, (2) sampling of participants, (3) description of participants and setting, (4) coverage of data analysis, (5) diagnostic methods, (6) reliability and standardization of measurements, and (7) response rate management. The reviewers rated each study using the options “yes,” “no,” and “unclear,” which were taken as “low,” “high,” and “unclear” risk of bias, respectively. The option “not applicable” was also available for each statement, each option had a score 1=yes, 0=no, 9=unclear. For the total score, the number of affirmative responses was summed, where more affirmative responses denoted lower risk of bias. The final quality score for each study was summed and classified as low (0 to 2 points), medium (3 to 4 points), or high (5 to 7 points; see Table S4). The first two authors independently performed the quality review of the included studies and agreed on a common score for each.

### 2.7. Data analyses

A descriptive summary and explanation of the characteristics and findings of all included studies were displayed in a comprehensive table.

Random effects meta-analyses of the reported proportions of college students with clinically significant symptoms of anxiety, depression and stress were carried out, and the mean prevalences with associated 95% confidence intervals (CI) were calculated. The three outcomes were analysed in two time periods: short-term adaptation (data collected in the first waves of Covid-19 between January and June 2020) and long-term (data collected from July 2020 to April 2022). *Q* and *Tau*^2^ statistics were computed to assess the statistical heterogeneity of effect sizes. Between-studies heterogeneity was also examined using the *Q* statistic, comparing the groups defined by the level of risk of bias, Covid wave, severity of confinement, geographical location, and measurement instrument used (1).

All analyses were carried out with the Stata statistical package (version SE 18) .

## 3. Results

### 3.1. Selection and inclusion of studies

The initial database search yielded a total of 1265 articles (see Figure 1). In the first phase after eliminating duplicates, 424 titles and abstracts were reviewed, of which 111 were selected for full-text evaluation with an inter-rater reliability of 92%. In the second phase after full-text screening, 23 studies were excluded with an inter-rater reliability of 91%, ten had objectives different from the review, five were non-empirical studies, two had data before COVID-19 and six do not include the target population. Finally, 90 studies were included in this systematic review, of which 74 reported sufficient data for meta-analysis.

**Figure 1.**
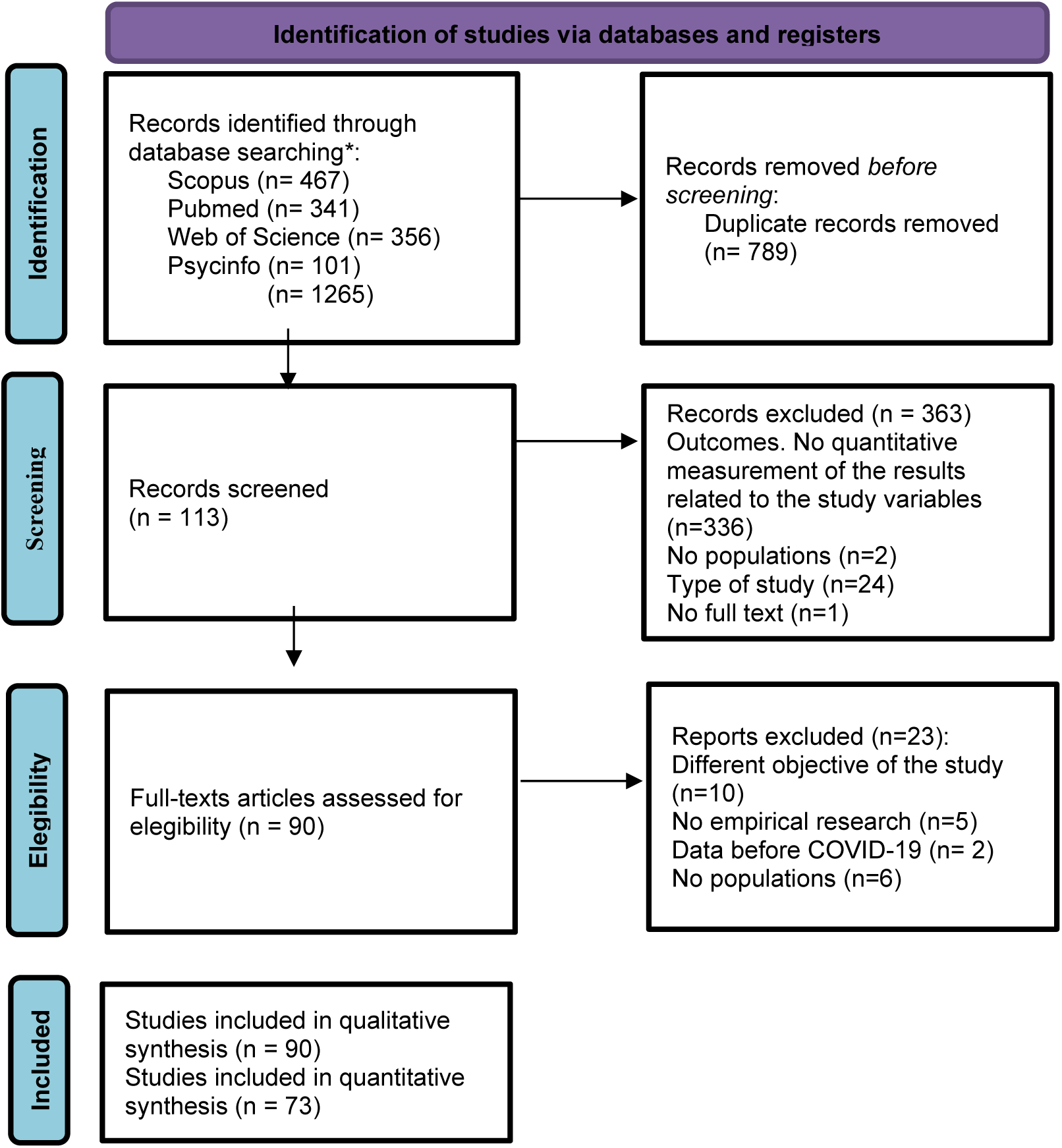
Flowchart of included articles.

### 3.2. Characteristics of the included studies

Of the 90 articles included in the review, 261,681 university students from 48 countries participated, most of the studies were conducted in China (11.4%); 65% of the participating population were women. Of note was a cross-national study that included 9 different nationalities (21). The age of the participants ranged from 18 to 78 years. Most studies (a total of 68) collected data in the first half of 2020 (January 1 to June 30, 2020; Supplementary Table S3).

### 3.3. Risk of bias of the selected studies

Table S4 shows the final quality assessment rates for each study. Following the JBI checklist for prevalence, 68 studies had scores of six and five, considering these studies as low risk of bias, 26 studies scored four, representing some concerns, and 1 study scored two (94), representing high risk of bias. The domain of sampling frame adequacy showed the lowest quality scores, followed by response rate management. This means that the included studies (1) did not adequately address the target population, presumably because they included unrepresentative samples or did not provide insight into the broader characteristics of the population, and (2) did not discuss information on how response rates were managed.

### 3.4. General outcomes (narrative and meta-analytic)

A total of 73 studies measured anxiety in 209,761 undergraduates; the most used instrument to measure anxiety was the GAD-7 (38 studies) and variants of the DASS (18 studies). All these studies were based on data collected through online surveys (100%). In relation to depression, 62 studies measured clinically significant symptoms during the past 15 days by means of variants of the PHQ (41 studies). Likewise, 36 studies measured stress, the most used questionnaire was the PSS in its different versions (21 studies). The DASS was the most used instrument in those studies that jointly assessed anxiety, depression, and stress (14 studies; see Table S3).

#### Mental health in short-term adaptation processes of university students during the COVID-19 pandemic

Short-term adaptation was assessed in the present study in the time interval from January to June 2020, coinciding with the first wave of the pandemic in all countries analysed [include reference to statistical source, e.g. the world in data] In total, 45 studies reported the prevalence of anxiety (22–66) among a total of 82,490 university students (Fig. 2; range, 1-93%), for depression a total of 35 studies reported prevalence (23–25,27,30,32,36–38,41–44,46–48,50,52–55,57,59,61–63,67–75) for a total of 61,318 college students (Fig. 3; range, 9-72%) and for stress 18 studies reported prevalence (23,24,30,38,42–44,46,47,50,50,54,55,57–59,61,76,77) for a total of 67,838 university students (Fig. 4; range, 22-87%). Regarding the use of assessment instruments for anxiety symptoms the GAD (27 studies) and variants of the SAS (12 studies) were the most used, for depression the most used scale was PHQ in all its variants (30 studies) and for stress the PSS in all its variants was the most used (15 studies), all studies assessed clinically significant symptoms over time, after one or two weeks, up to one month. One of these studies reported findings based on data from different months during this first survey period (63) but due to the overlap of survey months reported in the articles, these data were pooled. 100% of the articles were based on data collected online; this is applicable for all three variables studied. Regarding geographic location for both anxiety and depression the countries with the largest studies were China and Bangladesh, while for stress most of the data were collected in German population.

**Figure 2.**
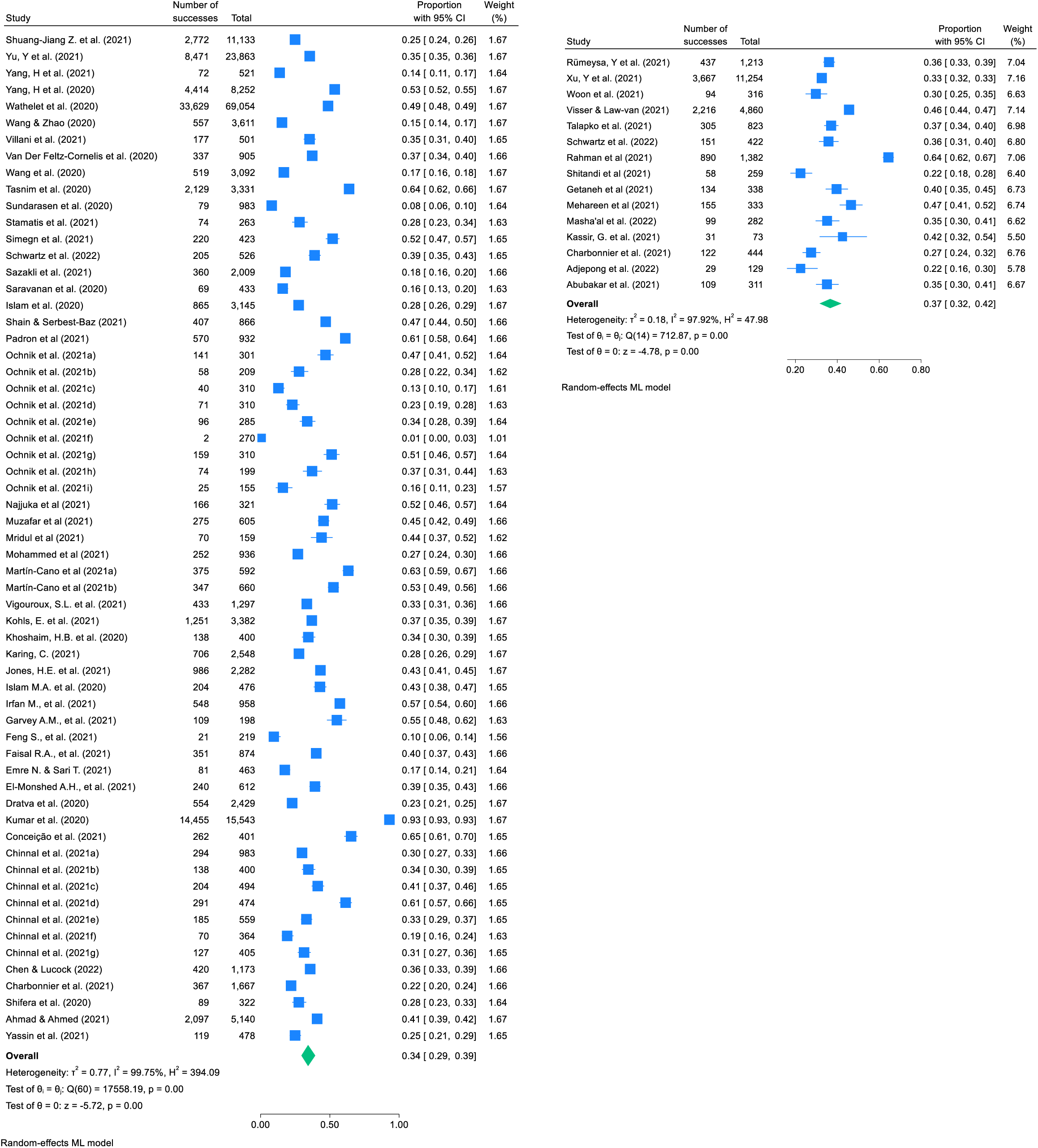
Pooled estimate of the proportion of college students with clinically significant symptoms of anxiety (Period 1 vs. Period 2)

**Figure 3.**
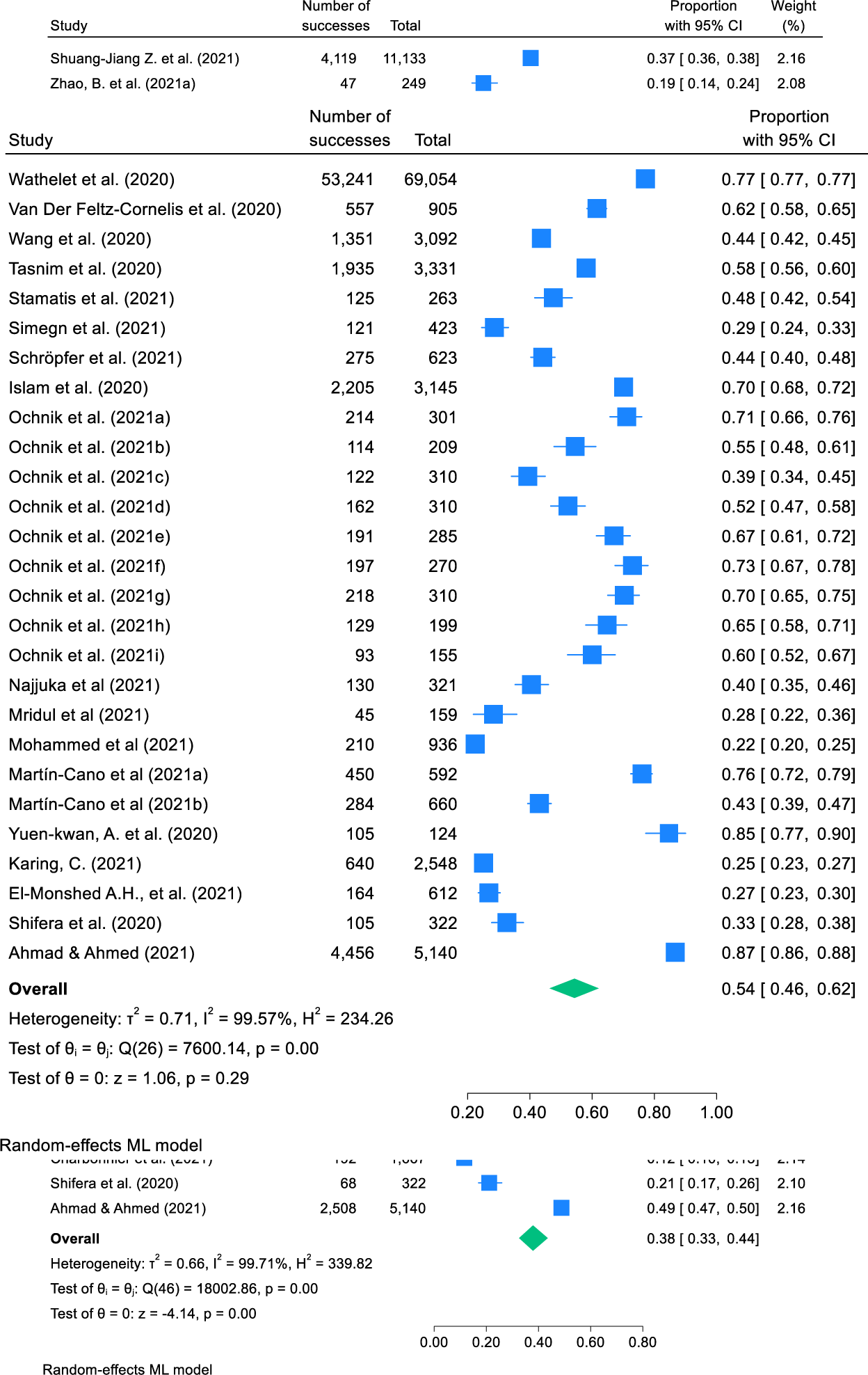
Pooled estimate of the proportion of college students with clinically significant symptoms of depression (period 1 vs period 2)

**Figure 4.**
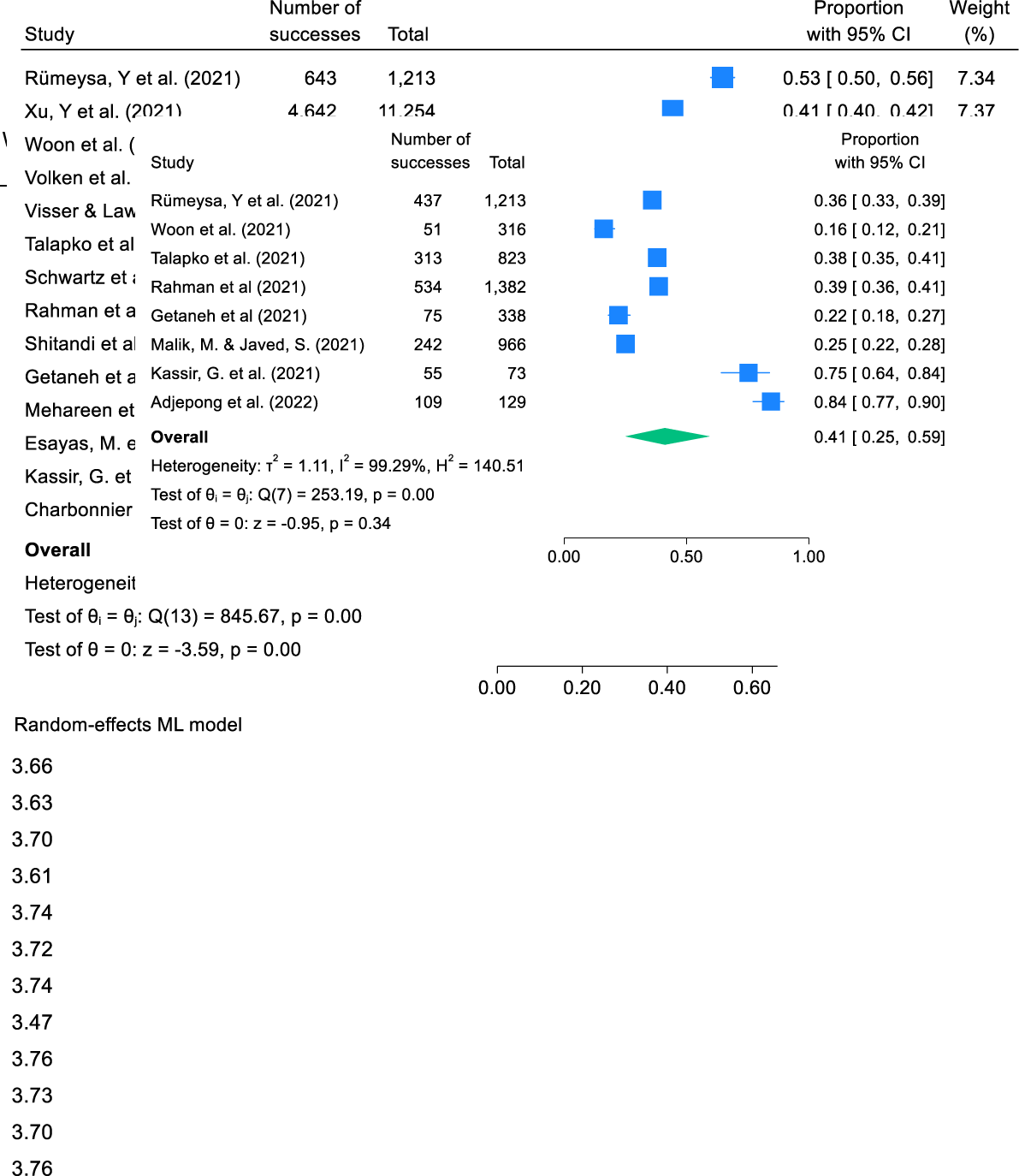
Pooled estimate of the proportion of college students with clinically significant symptoms of stress (period 1 vs period 2)

The estimated proportion of university students assessed with clinically significant symptoms of anxiety was 34% (95% confidence interval [CI] 29%-39%) with evidence of heterogeneity between studies (I^2^ = 99.75%), the prevalence of depression was 38% (95% confidence interval [CI] 33%-44%) with evidence of heterogeneity between studies (I^2^ = 99.71%); likewise, the prevalence for stress was 54% (95% confidence interval [CI] 46%-62%) with evidence of heterogeneity across studies (I^2^ =99.57%).

Heterogeneity tests showed that the effect values in each study were very heterogeneous and that there may have been significant regulatory variables. Therefore, a subgroup test was applied to explore sources of heterogeneity and regulation of effect size research characteristics, moderation analysis indicated that for anxiety and depression symptoms the severity of the pandemic, geographic location and measurement instrument were related, while for stress the COVID wave, geographic location and measurement instrument were moderators (Table 2). In the subgroup analysis, the highest prevalence was observed during the first wave of anxiety (95% CI 29%-38%) and stress (95% CI 45%-66%), the second wave of depression (95% CI 38%-48%), the highest levels of anxiety, depression and stress were observed in Portugal (65%), Bangladesh (64%) and United States - United Kingdom (84%), respectively. Studies using the STAI (n=1) and SAI (n=1) anxiety scales reported the highest pooled prevalence: 48% (95% CI: 48%-49%) and 47% (95% CI: 43%-50%), while studies using CAS (n=1) and HADS (n=3) reported the lowest pooled prevalence of anxiety of 15% (95% CI: 12%-19%), 23% (95% CI: 16%-33%) respectively, for depression, CES-D reported the highest prevalences of 72% (95% CI: 69%-75%) (Table S6). For stress, the PSS (either full or short version; n = 15) reported prevalences of 63 % (95 % CI: 53 %-72 %) (Table S5).

#### Mental health in long-term adjustment process of university students during the COVID-19 pandemic

For the second period corresponding to a long-term adaptation process, which runs from July 2020 to April 2022. A total of 14 studies were found that reported prevalences of anxiety (53,63,78–90), with a total of 8966 university students with clinically significant symptoms (Fig. 2; range 22%-64%). For that same time interval for depression a total of 9854 subjects reported clinically significant symptoms, corresponding to a total of 14 studies (Fig. 3; range 7%-73%) (53,61,63,79,79,82,84–92), for its part stress reported a total of 2245 university students with symptoms from 8 studies (Fig. 4; range 16%-84%) (63,81,82,84,86,87,89,93). The instruments most commonly used to collect prevalences of anxiety were the GAD (6 studies) and the DAS-21 (6 studies), for depression the PHQ-9 (6 studies) and for stress the DAS-21 (5 studies) all studies assessed clinically significant symptoms over time, over one to two weeks, up to one month.

The estimated proportion of college students assessed with clinically significant symptoms for the long-term period for anxiety was 37% (95% confidence interval [CI] 32%-42%) with significant evidence of heterogeneity across studies (I2 = 97.92%), for depression was 31% (95% confidence interval [CI] 23%-41%) with significant evidence of heterogeneity between studies (I2 = 99.49%) and for stress was 41% (95% confidence interval [CI] 25%-59%) with significant evidence of heterogeneity between studies (I2 = 99.29%).

In subgroup analysis, geographic location was related to symptoms of anxiety, depression, and stress, while instrument measurement was related to clinical symptoms of depression and stress (Table 3). The highest prevalences were observed during the first wave for the three study variables. The highest levels for anxiety, depression and stress were observed in Bangladesh (55%), Turkey (53%) and Lebanon (75%) respectively. Studies using the DASS 21 anxiety scales (n=6) reported the highest pooled prevalence: 40% (95 % CI: 30%-50%), for depression the highest prevalences were reported by the PHQ (either full or short version; n=9) with 29% (95 % CI: 22%-37%) and for stressor during quarantine (n=1) with prevalence of 75% (95 % CI: 64%-83%) (Table S6).

The prevalences assessed for the two periods are similar both in the short and long term for anxiety and depression, showing a slight increase of 3% in anxiety in the second period. period. With regard to depression and stress, a reduction of 7% and 13% respectively was found for the long-term assessment. Due to the overlapping of the confidence intervals, the differences between the two periods analysed are not statistically significant. It is also worth mentioning that the heterogeneity of results between studies is very large in all cases (I2 > 97%), as reflected in the Forest Plots.

## 4. Discussion

Our review found that there is no evidence of worsening mental health in the long-term period, which we interpret as the chronic phase, versus the short-term period coinciding with the first wave of the pandemic, understood as the acute stress/adaptive response. Although the confidence intervals are highly overlapping, there is a trend towards a slight reduction in stress (51% to 41%) and depression (38% to 31%), in anxiety there is a very slight rebound (34% to 37%) but this does not allow us to explain an increasing or decreasing evolution, we can simply infer that there is no worsening in mental health due to a cumulative effect. Results similar to those found in German university students with data obtained before and during the COVID-19 pandemic found no significant increase in stress and depression, only a minor elevation of anxiety between 2019 and 2020 (95), which is contradictory to other studies reporting high prevalence of anxiety and depression symptoms and stress in Swiss (96) and Italian (97) students. These results do not give evidence to the “pandemic fatigue”, a concept introduced by the World Health Organization (98) to explain a state of demotivation and tiredness influenced by emotions, experiences and perceptions related to exposure to a prolonged, severe, and restricted pandemic that sometimes leads to psychological discomfort and abandonment of prevention and self-care guidelines.

The results of the study also suggest a public health concern in relation to the high prevalences found in anxiety, depression and stress in university students when compared to other groups such as the general population, whose prevalences ranging from 15% to 33% in distress, anxiety, depression and stress (6,99,100), health workers 16% to 23% of distress (101,102), and older adults, with prevalences ranging from 12% to 28% (103,104). This confirms college students are a vulnerable group (105) explained to a large extent by an interaction of factors risks previous to the pandemic with factor risks intrinsic to the pandemic (i.e., more extensive restrictions than the rest of the population who recovered their routines much earlier than the university communities; 102,106,107). Students as a vulnerable group are exposed to new experiences and begin a stage of autonomy responding to family and personal expectations (108) that often exceeds their capacities and resources because they require greater self-regulation and emotional self-management (109), exposing themselves to psychosocial factors that can produce anxiety about the future, excessive academic load with long hours, exams, among others, all this generating the need to adapt to a new context.

The subgroup analysis for the present study allowed us to explore the prevalences for the first period and second period in relation to the confinement measures of being confined and leaving home with exceptions, as well as data collected during the first wave of COVID showed higher prevalences of anxiety, depression, and stress. For countries and territories, we found that Czechs (12%) and Nigerians (22%) for the first and second periods respectively were less anxious, as well as the number of new cases and deaths resulting from COVID-19 (110). The United States and the United Kingdom and Lebanon presented the highest levels of stress with 84%, explained by academic uncertainty, lack of reliable information, peer support (112) and financial uncertainty (113). The measures to mitigate COVID were also analysed regarding the the COVID wave, the hardiness of the confinement measures and the severity of the pandemic, observing high prevalences of anxiety and stress during the first wave, mobility restrictions and confinement measures due to misinformation regarding the virus, the increase in cases and deaths, and the lack of effective treatments to deal with the disease (114–116). It is important to highlight that, by including studies from different parts of the world, this can probably explain the heterogeneity of the data because mental health problems are culturally linked to how they are understood, conceptualized and evaluated (117), giving importance to the latter we found that the prevalences of distress varied according to the methodological differences for both the first and second period, in relation to the assessment instrument used in this case the highest grouped prevalence for anxiety of 48%, depression of 72% and stress of 63% was given by STAI, CES-D, PSS respectively for the first period and by the DASS, PHQ and Stressor during quarantine either in full or short version, were data similar to those found in other meta-analyses with general population (118) and university students (119) it should be noted that self-report measures are widely used and have been validated as a means to assess negative symptoms such as anxiety, depression and stress, because they allow spending less time and resources (120) but have several ways to be qualified, in relation to clinical ratings (severity ranges: low, medium, moderate or high) or absence and presence of symptoms which may explain the discrepancy between high and low score ranges, the selection of cut-off points represents a compromise between specificity and sensitivity according to the case used (121), in the present meta-analysis some studies do not report cut-off points (49).

### 4.1. Limitations and strengths

Our research should be interpreted considering limitations considering the high heterogeneity of the studies similar to data found in meta-analyses exploring mental health conditions in other populations during COVID-19 (122–124). In the present study, heterogeneity for anxiety (periods 1 and 2) and depression (periods 1 and 2) can be explained by factors intrinsic to the pandemic (severity in relation to restrictive measures) factors that may be both structural to the country and the country’s response to the pandemic (“geographic location” variable), but also due to different metrics employed (different instruments, with different versions, with different symptom severity classification cutoff points). In the present review we pooled data from different versions of the GAD, DAS, GHQ and PHQ measures, a practice that is common in systematic reviews but methodologically questionable (125), which is justified in the subgroup analysis where we found that prevalence estimates varied according to the measurement instrument used. On the other hand, it is important to highlight that our meta-analysis had a representative sample of college students with large sample sizes; however, most of the articles did not report in detail how response rates were handled. In addition, most of the studies are from Asia and Europe, few studies are from the West which could be interpreted in light of the data of interest, which was prevalence reporting, showing a gap in the literature from the Americas that needs to be addressed, limiting the degree of generalizability of the findings.

However, our study has important strengths, mainly complying with a rigorous protocol PRISMA guidelines, prospective registration in PROSPERO, validation of Boolean searches according to PRESS guidelines, the inclusion of the Rayyan tool in order to minimize the possible loss of evidence and the consensus review between two reviewers in the different phases of screening, data extraction and RoB. In turn, this review responds to proposals made by other authors (126) to provide greater support and scientific background to reviews related to the topic of COVID-19 and mental health in this case applied to students in a university context.

## 5. Conclusions

Our research provided an analysis of the high prevalence of anxiety, depression and stress in university students when compared with normal time and general population or other risk groups (health personnel, older adults, among others). This suggests that university students are a high-risk group. Furthermore, when comparing two periods of time for depression and stress, a slight decrease in prevalence is found, unlike anxiety, but this does not allow us to explain an increasing or decreasing evolution; we can simply infer that there is no worsening in mental health due to a cumulative effect.

It is necessary in view of the prevalence of mental health problems in university students to recognize the importance of prevention and promotion of mental health in Higher Education Institutions, approximately 56% of the students, although they present psychological discomfort do not consult, therefore, they do not receive attention from mental health professionals (127), among the factors of non-consultation is the inaccessibility to mental health services, negative attitudes and lack of knowledge about mental health problems (128). Educational institutions can play an important role in reducing stigma, increasing awareness about mental health services and creating a supportive environment on campus to encourage students to seek the help they need, as well as in the management of public policies at national levels with the Ministries of Education and/or Health that establish intervention and accompaniment strategies that promote more motivating, sustainable and healthy educational environments, being a strategic priority for Higher Education Institutions to prevent distress and improve wellbeing as a demand sought by the university student community.

## Data Availability

All data produced are available at:
https://figshare.com/articles/dataset/Dataset_Mental_health_and_individual_differences_in_the_short-_and_long-term_adaptation_processes_of_university_students_during_the_COVID-19_pandemic/21701228

https://figshare.com/articles/dataset/Dataset_Mental_health_and_individual_differences_in_the_short-_and_long-term_adaptation_processes_of_university_students_during_the_COVID-19_pandemic/21701228

## Data availability

Data are available at: https://figshare.com/articles/dataset/Dataset_Mental_health_and_individual_differences_in_the_short-_and_long-term_adaptation_processes_of_university_students_during_the_COVID-19_pandemic/21701228

## Authors’ contribution statement MPJV

study conceptualization, study design, meta-analysis data collection, data interpretation, writing: original draft, reviewing and editing. **JML:** study design, data analysis, writing: original draft, reviewing and editing. **LCV:** meta-analysis data collection, writing: review. **JPS:** writing: review. CM: writing: review. **AMR:** supervision, study design, writing: review and editing. **AS:** supervision, study design, writing: reviewing and editing. All authors contributed to and approved the final manuscript.

## PATIENT AND PUBLIC INVOLVEMENT

Patients and/or the public were not involved in the design, conduct, reporting, or dissemination plans of this research.

## PATIENT CONSENT FOR PUBLICATION

Not applicable.

## AI USE

Google Translate was used to check (and modify where necessary) the linguistic quality and suitability of the manuscript. No other AI tools were used in either the study or the writing of the manuscript.

---

**Table 1.** Logit transformed proportions.

|  | **Period 1** |  |  |  | **Period 2** |  |  |  |
| --- | --- | --- | --- | --- | --- | --- | --- | --- |
|  | Proportion | IC95% Li | IC95% Ls | *K* | Proportion | IC95% Li | IC95% Ls | *K* |
| **Anxiety** | 0.34 | 0.29 | 0.39 | 61 | 0.37 | 0.32 | 0.42 | 15 |
| **Depression** | 0.38 | 0.33 | 0.44 | 47 | 0.31 | 0.23 | 0.41 | 14 |
| **Stres** | 0.54 | 0.46 | 0.62 | 27 | 0.41 | 0.25 | 0.59 | 8 |

