## Supplementat file 1 for "Mental health in the short- and long-term adaptation processes of university students during the COVID-19 pandemic: A systematic review and meta-analysis"

**Supplementary Materials**

**Table S1.**

*Search Strategy of all Databases*

| |
|---|
| *Scopus:* ( TITLE ( pandemic ) OR TITLE ( covid ) OR KEY ( covid ) AND TITLE ( "mental health" ) OR TITLE ( depress* ) OR ABS ( depress* ) OR TITLE ( anxi* ) OR ABS ( anxi* ) OR TITLE ( stress ) OR ABS ( stress ) OR TITLE ( resilience* ) OR ABS ( resilience* ) OR TITLE ( wellbeing ) OR ABS ( wellbeing ) OR TITLE ( somati* ) OR ABS ( somati* ) OR TITLE ( "quality of life" ) OR ABS ( "quality of life" ) OR TITLE ( "prevent* behavio*" ) OR ABS ( "prevent* behavio*" ) OR TITLE ( personality ) OR ABS ( personality ) OR TITLE ( posttraumatic AND growth ) OR ABS ( posttraumatic AND growth ) OR TITLE ( loneliness ) OR ABS ( loneliness ) OR TITLE ( perceived AND control ) OR ABS ( perceived AND control ) AND TITLE ( students AND university ) ) |
| *Web Of Science:* (TI=(pandemic) OR TI=(COVID) OR AK=(COVID)) AND (TI=("mental health") OR TI=(depress*) OR AB=(depress*) OR TI=(anxi*) OR AB=(anxi*) OR TI=(stress) OR AB=(stress) OR TI=(resilience*) OR AB=(resilience*) OR TI=(wellbeing) OR AB=(wellbeing) OR TI=(somati*) OR AB=(somati*) OR TI=("quality of life") OR AB=("quality of life") OR TI=("prevent* behavio*") OR AB=("prevent* behavio*") OR TI=(posttraumatic growth) OR AB=(posttraumatic growth) OR TI=( perceived control ) OR AB=(perceived control) OR TI=(personality) OR AB=(personality))AND TI=(University students) |
| *PubMed:* (pandemic[Title] OR COVID[Title] OR COVID[Keywords]) AND ("mental health"[Title] OR depress*[Title/Abstract] OR anxi*[Title/Abstract] OR stress[Title/Abstract] OR resilience*[Title/Abstract] OR wellbeing[Title/Abstract] OR somati*[Title/Abstract] OR "quality of life"[Title/Abstract] OR "prevent* behavio*"[Title/Abstract] OR posttraumatic growth[Title/Abstract] OR perceived control[Title/Abstract] OR personality[Title/Abstract] AND University students [Title]) |
| *PsycInfo:* (ti(pandemic*) OR ti(COVID) OR if(COVID)) AND (ti("mental health") OR ti(depress*) OR ab(depress*) OR ti(Anxi*) OR ab(Anxi*) OR ti(Stress) OR ab(Stress) OR ti(resilience*) OR ab(resilience*) OR ti(wellbeing) OR ab(wellbeing) OR ti(somati*) OR ab(somati*) OR ti("quality of life") OR ab("quality of life") OR ti("prevent* behavio*") OR ab("prevent* behavio*") OR ti(posttraumatic growth) OR ab(posttraumatic growth) OR ti(perceived control) OR ab(perceived control) OR ti(Personality) OR ab(Personality)) AND ti(University students) |

**Table S2**

*Studies Included in the Systematic Review and Meta-Analysis (By Continent)*

| Author, Country | Title |
|---|---|
| **Asian Continent** | |
| Zhao et al., China and Korea | Assessing Knowledge, Preventive Practices, and Depression among Chinese University Students in Korea and China during the COVID-19 Pandemic: An Online Cross-Sectional Study |
| Zhao, et al., Korea, China and Japan | Novel Coronavirus (COVID-19) Knowledge, Precaution Practice, and Associated Depression Symptoms among University Students in Korea, China, and Japan. |
| Yu, et al., China | Factors Influencing Depression and Mental Distress Related to COVID-19 Among University Students in China: Online Cross-sectional Mediation Study. |
| Rümeysa, et al., Turkey | The Psychological Effects of COVID 19 on Medical and Non-medical University Students. |
| Yang, et al., China | Impact of coronavirus disease 2019 on the mental health of university students in Sichuan Province, China: An online cross-sectional study |
| Yang, et al., China | Opinions from the epicenter: an online survey of university students in Wuhan amidst the COVID-19 outbreak11 |
| Xu, et al., China | Prevalence and Risk Factors of Mental Health Symptoms and Suicidal Behavior Among University Students in Wuhan, China During the COVID-19 Pandemic |
| Woon et al., Malaysia | Depression, anxiety, and the COVID-19 pandemic: Severity of symptoms and associated factors among university students after the end of the movement lockdown. |
| Wang & Zhao, China | The Impact of COVID-19 on Anxiety in Chinese University Students. |
| Wang et al., China | Stress, anxiety, and sleep among college and university students during the covid-19 pandemic |
| Tasnim et al., Bangladesh | Suicidal ideation among Bangladeshi university students early during the COVID-19 pandemic: Prevalence estimates and correlates |
| Tang et al., China | Prevalence and correlates of PTSD and depressive symptoms one month after the outbreak of the COVID-19 epidemic in a sample of home-quarantined Chinese university students |
| Sundarasen et al., Malaysia | Psychological Impact of COVID-19 and Lockdown among University Students in Malaysia: Implications and Policy Recommendations. |

| | |
|---|---|
| Simegn et al., Ethiopia | Depression, anxiety, stress and their associated factors among Ethiopian University students during an early stage of COVID-19 pandemic: An online-based cross-sectional survey. |
| Saravanan et al., Arab Emirates | Knowledge, Anxiety, Fear, and Psychological Distress About COVID-19 Among University Students in the United Arab Emirates. |
| Islam et al., Bangladesh | Psychological responses during the COVID-19 outbreak among university students in Bangladesh |
| Shain & Serbest-Baz, Turkey | Views and Anxiety Levels of University Students Regarding Distance Education during the Covid-19 Pandemic |
| Rahman et al., Bangladesh | Depression, anxiety, and stress among public university students in Bangladesh during the COVID-19 pandemic |
| Pramukti et al., Indonesia, Taiwan and Thailand | Anxiety and Suicidal Thoughts During the COVID-19 Pandemic: Cross-Country Comparative Study Among Indonesian, Taiwanese, and Thai University Students. |
| Muzafar et al., Bangladesh | Generalized anxiety disorder among Bangladeshi university students during COVID-19 pandemic: gender specific findings from a cross-sectional study. |
| Mridul et al., India | Online Classes during COVID-19 Pandemic: Anxiety, Stress & Depression among University Students |
| Mohammed et al., Saudi Arabia | Psychological problems among the university students in Saudi Arabia during the COVID-19 pandemic. |
| Mehareen et al., Bangladesh | Prevalence and socio-demographic correlates of depression, anxiety, and co-morbidity during COVID-19: A cross-sectional study among public and private university students of Bangladesh |
| Masha'al et al., Jordan | Anxiety and coping strategies among nursing students returning to university during the COVID-19 pandemic. |
| Malik & Javed, Oman | Perceived stress among university students in Oman during COVID-19-induced e-learning |
| Khoshaim et al., Saudi Arabia | Anxiety Level of University Students During COVID-19 in Saudi Arabia. |
| Khan et al., China | Psychological Distress and Trust in University Management Among International Students During the COVID-19 Pandemic. |
| Kassir et al., Lebanon | Psychological distress experienced by self-quarantined undergraduate university students in lebanon during the covid-19 outbreak |
| Karmokar et al., Bangladesh | Depression and Behavioral Changes Associated with Social Media Dependency During COVID-19 Pandemic Among University Students in Bangladesh: A Cross-Sectional Study |

| | |
|---|---|
| Jing et al., China | Network-Based Online Survey Exploring Self-Reported Depression Among University and College Students During the Early Days of the COVID-19 Outbreak. |
| Islam et al., Bangladesh | Depression and anxiety among university students during the COVID-19 pandemic in Bangladesh: A web-based cross-sectional survey |
| Irfan et al., Malaysia | The Psychological Impact of Coronavirus on University Students and its Socio-Economic Determinants in Malaysia. |
| Feng et al., China | Fear and anxiety about covid-19 among local and overseas chinese university students |
| Faisal et al., Bangladesh | Mental health status, anxiety, and depression levels of bangladeshi university students during the covid-19 pandemic |
| Emre & Sari., Turkey | Evaluation of the Behavior, Anxiety and Stress of University Students in the New Type of Coronavirus Pandemic |
| El-Monshed et al., Egypt | University students under lockdown, the psychosocial effects and coping strategies during covid-19 pandemic: A cross sectional study in egypt |
| Durbas et al., Turkey | Anxiety and Stress Levels Associated With COVID-19 Pandemic of University Students in Turkey: A Year After the Pandemic. |
| Kumar et al., Bangladesh | Impact of COVID-19 on Psychology among the University Students |
| Chinnal et al., Malaysia, Saudi Arabia, Pakistan, Bangladesh, China, India, and Indonesia | Psychological impact of COVID-19 and lock down measures: An online cross-sectional multicounty study on Asian university students |
| Azmi et al., Saudi Arabia | Prevalence of COVID-19 Pandemic, Self-Esteem and Its Effect on Depression Among University Students in Saudi Arabia. |
| Ahmad & Ahmed., Saudi Arabia | The mental health impact of pandemic covid-19 crisis on university students in saudi arabia and associated factors |
| Abuhmaidan & Al-Majali., Arab Emirates | The impact of the coronavirus pandemic on mental health among al ain university students in light of some demographic variables |
| Abubakar et al., Indonesia | Anxiety to COVID-19 pandemic amongst university students is related with gastrointestinal symptoms |
| **European Continent** | |
| Zurlo et al., Italia | Psychological Health Conditions and COVID-19-Related Stressors Among University Students: A Repeated Cross-Sectional Survey |
| Werner et al., Germany | The impact of lockdown stress and loneliness during the COVID-19 pandemic on mental health among university students in Germany. |

| | |
|---|---|
| Wathelet et al., Francia | Factors Associated with Mental Health Disorders Among University Students in France Confined During the COVID-19 Pandemic. |
| Volken et al., Switzerland | Depressive Symptoms in Swiss University Students during the COVID-19 Pandemic and Its Correlates. |
| Villani et al., Italia | Impact of the COVID-19 pandemic on psychological well-being of students in an Italian university: a web-based cross-sectional survey. |
| Van Der Feltz-Cornelis et al., England | Workplace Stress, Presenteeism, Absenteeism, and Resilience Amongst University Staff and Students in the COVID-19 Lockdown. |
| Talapko et al., Croacia | Mental Health and Physical Activity in Health-Related University Students during the COVID-19 Pandemic. |
| Spatafora et al., Germany | Fear of Infection and Depressive Symptoms among German University Students during the COVID-19 Pandemic: Results of COVID-19 International Student Well-Being Study. |
| Schröpfer et al., Germany | Psychological Stress among Students in Health-Related Fields during the COVID-19 Pandemic: Results of a Cross-Sectional Study at Selected Munich Universities. |
| Sazakli et al., Greece | Prevalence and associated factors of anxiety and depression in students at a Greek university during COVID-19 lockdown. |
| Rogowska et al., Poland | Changes in mental health during three waves of the COVID-19 pandemic: A repeated cross-sectional study among Polish university students |
| Rogowska et al., Poland | Examining Anxiety, Life Satisfaction, General Health, Stress and Coping Styles During COVID-19 Pandemic in Polish Sample of University Students. |
| Padron et al., Spain | A Study on the Psychological Wound of COVID-19 in University Students |
| Matos-Fialho et al., Germany | Perceptions of Study Conditions and Depressive Symptoms During the COVID-19 Pandemic Among University Students in Germany: Results of the International COVID-19 Student Well-Being Study |
| Marcén-Román et al., Spain | Stress Perceived by University Health Sciences Students, 1 Year after COVID-19 Pandemic |
| Vigouroux et al., France | The psychological vulnerability of French university students to the COVID-19 confinement |
| Kostić et al., Serbia | Perceived stressÂ among university students in south-east Serbia during the COVID-19 outbreak. |
| Kohls et al., Germany | Mental Health, Social and Emotional Well-Being, and Perceived Burdens of University Students During COVID-19 Pandemic Lockdown in Germany. |
| Karing, Germany | Prevalence and predictors of anxiety, depression and stress among university students during the period of the first lockdown in Germany. |
| Garvey et al., Spain | The Psychological Impact of Strict and Prolonged Confinement on Business Students during the COVID-19 Pandemic at a Spanish University. |

| | |
|---|---|
| Dratva, Switzerland | Swiss University Students' Risk Perception and General Anxiety during the COVID-19 Pandemic. |
| Di Consiglio et al., Italia | The Impact of COVID-19 Pandemic on Italian University Students' Mental Health: Changes across the Waves |
| Conceição et al., Portugal | The Association Between Changes in the University Educational Setting and Peer Relationships: Effects in Students' Depressive Symptoms During the COVID-19 Pandemic |
| Chen & Lucock, United Kingdom | The mental health of university students during the COVID-19 pandemic: An online survey in the UK. |
| Charbonnier et al., France | Psychological Vulnerability of French University Students during the COVID-19 Pandemic: A Four-Wave Longitudinal Survey. |
| Brailovskaia et al., Germany | Suicide ideation during the COVID-19 outbreak in German university students: Comparison with pre-COVID 19 rates. |
| Arënliua et al., Kosovo | Anxiety and depression among Kosovar university students during the initial phase of outbreak and lockdown of COVID-19 pandemic |
| **Africa Continent** | |
| Visser & Law-va, South Africa | University students' mental health and emotional wellbeing during the COVID-19 pandemic and ensuing lockdown |
| Shitandi et al., Nigeria | Impact of Covid-19 on the Mental Health of Delta State University students, Nigeria. |
| Najjuka et al., Ugandan | Depression, anxiety, and stress among Ugandan university students during the COVID-19 lockdown: An online survey |
| Getaneh et al., Ethiopia | The Psychological Impact of COVID-19 Pandemic on Graduating Class Students at the University of Gondar, Northwest Ethiopia. |
| Esayas, et al., Ethiopia | Prevalence and Associated Factors of Depressive Symptoms Among Mizan-Tepi University Students During the COVID-19 Pandemic. |
| Shifera et al., Ethiopia | The Psychological Impacts of COVID-19 Pandemic Among University Students in Bench-Sheko Zone, South-west Ethiopia: A Community-based Cross-sectional Study. |
| Assefa et al., Ethiopia | Mental Health Disorders During COVID-19 Pandemic Among Southwest Ethiopia University Students: An Institutional-Based Cross-Sectional Study. |
| Adjepong et al., Ghana | Limited negative effects of the COVID-19 pandemic on mental health measures of Ghanaian university students. |
| Yassin et al., Sudanese | Anxiety among the Sudanese university students during the initial stage of COVID-19 pandemic. |
| **American Continent** | |

| | |
|---|---|
| Stamatis et al., United States | A longitudinal investigation of COVID-19 pandemic experiences and mental health among university students |
| Schwartz et al., United States | Mental and physical health among students at a private university that held in-person classes during the covid-19 pandemic |
| Rodríguez-Hidalgo et al., Ecuador | Fear of COVID-19, Stress, and Anxiety in University Undergraduate Students: A Predictive Model for Depression |
| Jones et al., New York | The Impact of the COVID-19 Pandemic on College Students' Health and Financial Stability in New York City: Findings from a Population-Based Sample of City University of New York (CUNY) Students. |
| García-Espinosa et al., Mexican | Psychosocial impact on health-related and non-health related university students during the COVID-19 pandemic. Results of an electronic survey. |
| **Oceania Continent** | |
| Liu et al., Australia | Addressing Depression Symptoms among University Students under COVID-19 Restrictions-The Mediating Role of Stress and the Moderating Role of Resilience. |
| **European and American Continent** | |
| Martín-Cano et al., Spain and Mexican | Depression, anxiety and stress in social work students during covid-19 confinement. A comparative study of spanish and mexican universities |
| Yuen-kwan et al., United Kingdom and United Stated | Mental Health Impacts of the COVID-19 Pandemic on International University Students, Related Stressors, and Coping Strategies. |
| **Asia and European Continent** | |
| Herbert et al., Egypt and Germany | How do you feel during the COVID-19 pandemic? A survey using psychological and linguistic self-report measures, and machine learning to investigate mental health, subjective experience, personality, and behaviour during the COVID-19 pandemic among university |
| **Asia, European and American Continent** | |
| Ochnik et al., Poland, Slovenia, Czechia, Ukraine, Russia, Germany, Turkey, Israel and Colombia | Mental health prevalence and predictors among university students in nine countries during the COVID-19 pandemic: a cross-national study. |

**Table S3**

*Main Characteristics of the Included Studies (k = 90; N = 261.681)*

| Author, Year | Country | Females (%) | Data collection date | Sample size (N) | $M_{age}$ (SD) | Anxiety | | | Depression | | | Stress | | | |
|---|---|---|---|---|---|---|---|---|---|---|---|---|---|---|---|
| | | | | | | Assessment tool | Prevalence (%) | Mean (SD) | Assessment tool | Prevalence (%) | Mean (SD) | Assessment tool | Prevalence (%) | Mean (SD) | Ass |
| Zurlo, M.C. et al. (2022) | Italia | 75.4 | 1/4/2020 | 197 | 21.3 (3.27) | SCL-90-R - ANX | NR | 1.03 (0.70) | SCL-90-R DEP | NR | 1.24 (0.71) | CSSQ | NR | 10.49 (4.32) | |
| | Italia | 75.4 | 1/11/2020 | 274 | 21.3 (3.27) | SCL-90-R - ANX | NR | 1.16 (0.75) | SCL-90-R DEP | NR | 1.49 (0.83) | CSSQ | NR | 13.51 (4.52) | |
| | Italia | 75.4 | 1/4/2021 | 200 | 21.3 (3.27) | SCL-90-R - ANX | NR | 1.44 (0.82) | SCL-90-R DEP | NR | 1.71 (0.83) | CSSQ | NR | 14.01 (4.85) | |
| Shuang-Jiang Z. et al. (2021) | China | 62.3 | 1/3/2020 | 11133 | ≥18-35 | GAD-7 | 24.9 Anxiety; 75.1 No anxiety | NR | PHQ-9 | 37 Depression; 63 No Depression | NR | NR | NR | NR | |
| Zhao, B. et al. (2021) | China | 69.48 | 1/3/2020 | 249 | 22.12 (2.28) | NR | NR | NR | PHQ-9 | 18.9 Depression; 81.1 No Depression | 6.20 (0.31) | NR | NR | NR | |
| | Korea | 66.67 | | 171 | 24.08 (4.14) | NR | NR | NR | PHQ-9 | 28.7 Depression; 71.3 No Depression | 7.20 (0.41) | NR | NR | NR | |
| Zhao, B. et al. (2020) | Korea | 58.4 | 20/3/2020 | 390 | 23.14 (0.15) | NR | NR | NR | PHQ-9 | 49.23 Depression; 50.77 No Depression | 5.94 (5.44) | NR | NR | NR | |
| | China | 70.4 | | 281 | 23.63 (0.18) | NR | NR | NR | PHQ-9 | 60.5 Depression; 39.50 No Depression | 6.40 (5.12) | NR | NR | NR | |
| | Japan | 60 | | 150 | 24.13 (0.53) | NR | NR | NR | PHQ-9 | 60 Depression; 40 No Depression | 7.33 (6.20) | NR | NR | NR | |

| | | | | | | | | | | | | | | |
|---|---|---|---|---|---|---|---|---|---|---|---|---|---|---|
| Yu, Y et al. (2021) | China | 68.1 | 10/2/2020 | 23863 | NA | Mental Distress Due to COVID-19 | 35.5 Anxiety; 64.5 no anxiety | NR | PHQ-9 | 39.1 Depression; 60.9 No Depression | NR (5.50) | NR | NR | NR |
| Rümeysa, Y et al. (2021) | Turkey | 73 | 20/10/2020 | 1213 | 20.74 (3.32) | DASS - 21 | 51 Anxiety; 49 No anxiety | 4.80 (4.37) | DASS - 21 | 67 Depression; 34 No Depression | 8.12 (5.89) | DASS -21 | 49 Stress; 51 No stress | 08.08 (5.33) |
| Yang, H et al. (2021) | China | 77.5 | 1/4/2020 | 521 | 22.02 (1.76) | SAS | 45.3 Anxiety; 54.7 No anxiety | NR | NR | NR | NR | NR | NR | NR |
| Yang, H et al. (2020) | China | 61.25 | 1/1/2020 | 8252 | NA | NR | 53.49 Anxiety; 46.51 No anxiety | 3.53 (0.88) | NR | NR | NR | NR | NR | NR |
| Xu, Y et al. (2021) | China | NR | 1/6/2020 | 11254 | NA | GAD-7 | 32.58 Anxiety; 67.42 No anxiety | NR | PHQ-9 | 41.52 Depression; 58.48 No Depression | NR | NR | NR | NR |
| Woon et al. (2021) | Malaysia | 69.9 | 1/7/2020 | 316 | 21(NA) | DASS-21 | 36.7 Anxiety; 63.3 No anxiety | 4.28 (NR) | DASS-21 | 36.4 Depression; 63.6 No Depression | 4.61 (NR) | DASS-21 | 42.4 Stress; 57.6 No stress | 5.34 (NR) |
| Werner et al. (2021) | Germany | 72.6 | 1/6/2020 | 3066 | NA | GAD-2 | NR | 2.12 (1.73) | PHQ-9 | NR | 8.49 (5.31) | NR | NR | NR |
| Wathelet et al. (2020) | Francia | 72.8 | 17/4/2020 | 69054 | ≥18-22 | STAI Y-2 | 22.44 Anxiety; 77.56 No anxiety | 45 (NR) | BDI-13 | 74.4 Depression; 25.5 No Depression | 7 (NR) | PSS-10 | 77.1 Stress; 22.8 No stress | 20 (NR) |

| | | | | | | | | | | | | | | |
|---|---|---|---|---|---|---|---|---|---|---|---|---|---|---|
| Wang & Zhao (2020) | China | 59.7 | 15/02/2020 | 3611 | ≥18-24 | SAS | 15.43 Anxiety; 84.57 No anxiety | 40.53 (10.15) | NR | NR | NR | NR | NR | NR |
| Volken et al. (2021) | Switzerland | 69.8 | 14/04/2020 | 2363 | 26.4 (5.6) | NR | 27.1 Anxiety; 72.9 No anxiety | 7.3 (4.7) | PHQ-9 | NR | NR | NR | NR | NR |
| Volken et al. (2021) | Switzerland | 69 | 13/10/2020 | 1208 | 25 (5.4) | NR | 27.7 Anxiety; 72.9 No anxiety | 7.6 (5.0) | PHQ-9 | NR | NR | NR | NR | NR |
| Visser & Law-van (2021) | South Africa | 66.4 | 01/07/2020 | 4860 | NA | GAD-2 | 45.6 Anxiety; 54.4 No anxiety | 1.69 (1.37) | PHQ-2 | 35 Depression; 65 No Depression | 1.91 (1.24) | NR | NR | NR |
| Villani et al. (2021) | Italia | 71.4 | 08/06/2020 | 501 | NA | SAS | 35.3 Anxiety; 64.7 No anxiety | NR | SDS | 72.9 Depression; 27,1 No depression | NR | NR | NR | NR |
| Van Der Feltz-Cornelis et al. (2020) | England | 72 | 13/05/2020 | 905 | 27.5 (NA) | GAD- 7 | 37.2 Anxiety; 62.8 No anxiety | 8.31 (5.74) | PHQ-9 | 46.5 Depression; 53.5 No Depression | 9.87 (6.57) | PSQ | 61.5 Stress; 38.5 No stress | 0.51 (0.20) |
| Wang et al. (2020) | China | 66.4 | 21/02/2020 | 3092 | NA | GAD-7 | 16.8 Anxiety; 83.2 No anxiety | NR | NR | NR | NR | PSS-10 | 43.7 Stress; 56.3 No stress | NR |
| Tasnim et al. (2020) | Bangladesh | 40.6 | 01/04/2020 | 3331 | 21.4 (1.9) | DASS-21 | 63.9 Anxiety; 36.1 No anxiety | NR | DASS-21 | 63 Depression; 37 No Depression | NR | DASS-21 | 58.1 Stress; 41.1 No stress | NR |

| | | | | | | | | | | | | | | |
|---|---|---|---|---|---|---|---|---|---|---|---|---|---|---|
| Tang et al. (2020) | China | 60.8 | 20/02/2020 | 2485 | 19.8 (1.5) | NR | NR | NR | PHQ-9 | 9 Depression; 91 No depression | NR | NR | NR | NR |
| Talapko et al. (2021) | Croacia | 81.4 | 26/11/2020 | 823 | 29 (NA) | DASS-21 | 50.9 Anxiety; 49.1 No anxiety | 5.18 (5.20) | DASS-21 | 50.8 Depression; 49.2 No Depression | 06.01 (5.35) | DASS-21 | 49.9 Stress; 50.1 No stress | 8.12 (5.84) |
| Sundarasen et al. (2020) | Malaysia | 66.4 | 20/04/2020 | 983 | NA | SAS | 8 Anxiety; 92 No anxiety | NR | NR | NR | NR | NR | NR | NR |
| Stamatis et al. (2021) | United States | 67.7 | 01/03/2020 | 165 | 19.55 (1.7) | GAD-7 | 28.5 Anxiety; 71.5 No anxiety | 6.68 (6.17) | PHQ-9 | 35.5 Depression; 65.5 No Depression | 8.44 (6.98) | IES-6 | 50.3 Stress; 49.7 No stress | 1.82 (01.01) |
| | United States | 67.7 | 01/05/2020 | 98 | 19.55 (1.7) | GAD-7 | 27.6 Anxiety; 72.4 No anxiety | 7.20 (6.62) | PHQ-9 | 43.9 Depression; 56.1 No Depression | 9.36 (7.30) | IES-6 | 42.9 Stress; 57.1 No stress | 1.62 (0.99) |
| Spatafora et al. (2022) | Germany | 70 | 01/05/2020 | 5021 | 24.4 (4.5) | NR | NR | NR | CES-D 8 | NR | 9.25 (4.7) | NR | NR | NR |
| Simegn et al. (2021) | Ethiopia | 35.7 | 30/06/2020 | 423 | 22.96 (NA) | DASS-21 | 52 Anxious - 48.0 No anxious | NR | DASS-21 | 46.3 Depression; 53.7 NO Depression | NR | DASS-21 | 28.7 Stress; 71.4 No stress | NR |
| Schwartz et al. (2022) | United States | 74 | 19/05/2020 | 526 | 20.6 (NA) | PHQ4 | 39 Anxiety; 61 No anxiety | NR | PHQ4 | 35.9 Depression; 64.1 No depression | NR | NR | NR | NR |

| | | | | | | | | | | | | | | |
|---|---|---|---|---|---|---|---|---|---|---|---|---|---|---|
| | United States | NR | 10/11/2020 | 422 | 20.6 (NA) | PHQ4 | 50.8 Anxiety; 49.2 No anxiety | NR | PHQ4 | 33.5 Depression; 66.5 No Depression | NR | NR | NR | NR |
| Schröpfer et al. (2021) | Germany | 82.5 | 29/06/2020 | 623 | NA | NR | NR | NR | NR | NR | NR | BRCS | 44.1 Stress; 55.8 No stress | NR |
| Sazakli et al. (2021) | Greece | NR | 15/04/2020 | 2009 | 22.0 (3.4) | HADS-A | 35.8 Anxiety; 64.2 No anxiety | 6.6 (4.4) | HADS-D | 51.2 Depression; 48.8 No Depression | 8.5 (4.2) | NR | NR | NR |
| Saravanan et al. (2020) | Arab Emirates | 35.8 | 01/05/2020 | 433 | 21 (2.9) | CAS | 15.9 Anxiety; 84.1 No anxiety | 5.4 (3.6) | NR | NR | NR | NR | NR | NR |
| Islam et al. (2020) | Bangladesh | 40.5 | 01/04/2020 | 3145 | 21.4 (2.0) | DASS-21 | 27.5 Anxiety; 72.5 No anxiety | NR | DASS-21 | 76.1 Depression; 23.9 No Depression | NR | DASS-21 | 70.1 Stress; 29.9 No stress | NR |
| Shain & Serbest-Baz (2021) | Turkey | 78.2 | 01/06/2020 | 866 | 20 (06.04) | SAI | 100 Anxiety | 59 (29) | NR | NR | NR | NR | NR | NR |
| Rogowska et al. (2021) | Poland | 60.43 | 30/03/2020 | 657 | 23.68 (2.66) | GAD-7 | NR | 8.39 (5.63) | NR | NR | NR | PSS-10 | NR | 28.72 (5.70) |
| | Poland | 51.99 | 03/11/2020 | 651 | 22.72 (3.15) | GAD-7 | NR | 7.32 (5.64) | NR | NR | NR | PSS-10 | NR | 23.15 (05.07) |
| | Poland | 61.51 | 10/04/2021 | 634 | 23.31 (3.51) | GAD-7 | NR | 9.37 (5.94) | NR | NR | NR | PSS-10 | NR | 23.53 (4.76) |
| Rogowska et al. (2020) | Poland | 43.11 | 30/03/2020 | 914 | 23.04 (2.60) | GAD-7 | 65 Anxiety; 35 No anxiety | 7.85 (5.59) | NR | NR | NR | PSS | 56 Stress; 44 No stress | 20.32 (8.38) |

| | | | | | | | | | | | | | | |
|---|---|---|---|---|---|---|---|---|---|---|---|---|---|---|
| Rodríguez-Hidalgo et al. (2020) | Ecuador | 72 | NR | 640 | 21.69 (04.09) | DASS-21 | NR | 5.53 (4.98) | DASS-21 | NR | 5.93 (05.07) | DASS-21 | NR | 6.89 (5.54) |
| Rahman et al. (2021) | Bangladesh | 53.04 | 01/09/2020 | 1382 | ≥18-35 | DASS-21 | 71.56 Anxiety; 28.44 No anxiety | NR | DASS-21 | 74.53 Depression; 25.47 No Depression | NR | DASS-21 | 53.47 Stress; 46.53 No stress | NR |
| | Indonesia | 85.7 | 10/04/2020 | 938 | NA | STAI | NR | 2.33 (0.48) | NR | NR | NR | NR | NR | NR |
| Pramukti et al. (2020) | Taiwan | 62.6 | 10/04/2020 | 734 | NA | STAI | NR | 2.08 (0.42) | NR | NR | NR | NR | NR | NR |
| | Thailand | 80.1 | 10/04/2020 | 313 | NA | STAI | NR | 2.55 (0.43) | NR | NR | NR | NR | NR | NR |
| Padron et al. (2021) | Spain | 75.5 | 27/04/2020 | 932 | NA | GAD-7 | 61.2 Anxiety; 38.8 No anxiety | 10.82 (5.00) | PHQ-9 | 65.8 Depression; 34.2 No Depression | 12.98 (6.46) | NR | NR | NR |
| Shitandi et al. (2021) | Nigeria | 54.4 | 01/07/2020 | 259 | 21.50 (02.04) | GAD-7 | 52.1 Anxiety; 47.9 No anxiety | 6.0 (5.37) | PHQ-9 | 55.6 Depression; 44.4 No Depression | 07.07 (6.36) | NR | NR | NR |
| | Poland | 73.40 | 19/05/2020 | 301 | NA | GAD-7 | 76.5 Anxiety; 23.60 No anxiety | 9.20 (5.49) | PHQ-8 | 49.20 Depression; 50.8 No Depression | 9.90 (6.08) | PSS-10 | 92 Stress; 8 No stress | 22.69 (6.33) |
| Ochnik et al. (2021) | Slovenia | 85.20 | 14/05/2020 | 209 | NA | GAD-7 | 66 Anxiety; 34 No anxiety | 7.37 (5.33) | PHQ-8 | 35.50 Depression; 64.5 No Depression | 7.98 (6.27) | PSS-10 | 80.3 Stress; 19.60 No stress | 19.83 (7.56) |
| | Czechia | 65.80 | 17/06/2020 | 310 | NA | GAD-7 | 47.1 Anxiett; 59.2 No anxiety | 4.86 (3.98) | PHQ-8 | 21 Depression; 79 No Depression | 5.90 (4.70) | PSS-10 | 84.9 Stress; 15.20 No stress | 18.16 (3.99) |

| | | | | | | | | | | | | | | |
|---|---|---|---|---|---|---|---|---|---|---|---|---|---|---|
| | Ukraine | 70 | 14/05/2020 | 310 | NA | GAD-7 | 54.2 Anxiety; 45.8 No anxiety | 6.15 (5.11) | PHQ-8 | 20 Depression; 71 No Depression | 7.32 (5.39) | PSS-10 | 84.9 Stress; 15.20 No stress | 19.93 (5.99) |
| | Russia | 67 | 01/06/2020 | 285 | NA | GAD-7 | 63.9 Anxiety; 36.1 No anxiety | 7.48 (5.70) | PHQ-8 | 41.10 Depression; 58.9 No Depression | 9.00 (6.41) | PSS-10 | 88.8 Stress; 11.20 No stress | 21.98 (6.95) |
| | Germany | 71.50 | 02/06/2020 | 270 | NA | GAD-7 | 25.2 Anxiety; 74.8 No anxiety | 2.92 (3.26) | PHQ-8 | 37.70 Depression; 62.2 No Depression | 8.42 (4.89) | PSS-10 | 97.1 Stress; 3 No stress | 22.54 (05.05) |
| | Turkey | 55.80 | 16/05/2020 | 310 | NA | GAD-7 | 87.1 Anxiety; 12.9 No anxiety | 10.41 (5.25) | PHQ-8 | 62.30 Depression; 37.7 No Depression | 12.42 (06.02) | PSS-10 | 93.5 Stress; 6.50 No stress | 22.71 (6.43) |
| | Israel | 74.90 | 21/05/2020 | 199 | NA | GAD-7 | 67.9 Anxiety; 32.30 No anxiety | 7.92 (5.56) | PHQ-8 | 42.20 Depression;57.8 No Depression | 8.94 (5.81) | PSS-10 | 87.9 Stress; 12.10 No stress | 21.51 (6.88) |
| | Colombia | 65.20 | 05/06/2020 | 155 | NA | GAD-7 | 69 Anxiety; 31 No anxiety | 8.45 (5.79) | PHQ-8 | 49.70 Depression; 50.3 No Depression | 10.13 (6.41) | PSS-10 | 82.6 Stress; 17.40 No stress | 21.37 (7.55) |
| Najjuka et al. (2021) | Ugandan | 38.3 | 29/06/2020 | 321 | 24.8 (5.1) | DASS-21 | 98.4 Anxiety; 1.6 No anxiety | NR | DASS-21 | 80.7 Depression; 19.3 No Depression | NR | DASS-21 | 77.9 Stress; 22.1 No stress | NR |
| Muzafar et al. (2021) | Bangladesh | 59.5 | 01/05/2020 | 605 | 23.1 (3.4) | GAD-7 | 87.43 Anxiety ; 12.5 No anxiety | 9.6 (4.6) | NR | NR | NR | NR | NR | NR |

| | | | | | | | | | | | | | | |
|---|---|---|---|---|---|---|---|---|---|---|---|---|---|---|
| Mridul et al. (2021) | India | NR | 09/07/2020 | 159 | NA | DASS-21 | 56.58 Anxiety; 43.39 No anxiety | NR | DASS-21 | 50.32 Depression; 49.69 No Depression | NR | DASS-21 | 37.11 Stress; 62.89 No stress | NR |
| Mohammed et al. (2021) | Saudi Arabia | 56.4 | 04/06/2020 | 936 | NA | DASS-21 | 26.9 Anxiety; 73.1 No anxiety | NR | DASS-21 | 41.1 Depression; 58.9 No Depression | NR | DASS-21 | 22.4 Stress; 77.6 No stress | NR |
| Getaneh et al. (2021) | Ethiopia | 43.8 | 10/11/2020 | 338 | 24.70 (2.78) | DASS-21 | 39.60 Anxiety; 60.4 No anxiety | NR | DASS-21 | 40.20 Depression; 59.8 No Depression | NR | DASS-21 | 22.20 Stress; 77.8 No stress | NR |
| Mehareen et al. (2021) | Bangladesh | 51.95 | 18/07/2020 | 333 | 21.49 (1.56) | GAD-7 | 75.7 Anxiety; 24.30 No anxiety | NR | PHQ-9 | 82.3 Depression; 17.7 No Depression | NR | NR | NR | NR |
| Matos-Fialho et al. (2021)ç | Germany | 69 | 19/05/2020 | 5021 | 24 (5.1) | | NR | NR | CES-D8 | NR | 9.25 (0.67) | NR | NR | NR |
| Masha'al et al. (2022) | Jordan | 74.1 | 19/07/2020 | 282 | 20.08 (01.08) | GAD-7 | 70.6 Anxiety; 29.4 No anxiety | NR | NR | NR | NR | NR | NR | NR |
| Martín-Cano et al. (2021) | Spain | 90.4 | 01/05/2020 | 592 | 23.16 (5.78) | DASS-21A | 63.3 Anxiety; 36.7 No anxiety | NR | DASS-21D | 71.6 Depression; 28.4 No Depression | NR | DASS-21S | 76 Stress; 24 No stress | NR |

| | | | | | | | | | | | | | | | |
|---|---|---|---|---|---|---|---|---|---|---|---|---|---|---|---|
| | Mexican | 94.2 | 01/05/2020 | 660 | 21.81 (4.77) | DASS-21A | 52.6 No Anxiety; 47.4 Anxiety | NR | DASS-21D | 47.6 Depression; 52.4 No Depression | NR | DASS-21S | 43 Stress; 57 No stress | NR | |
| Marcén-Román, Y. et al. (2021) | Spain | 81.7 | 01-2021 | 252 | 21.02 (5.02) | GADS | 71.4 Anxiety; 28.6 No anxiety | NR | GADS | 81 Depression; 19 No Depression | NR | PSS-10-C | 13.1 Stress; 86.9 No stress | NR | |
| Malik, M. & Javed, S. (2021) | Oman | 84.4 | 12-2020 | 966 | NA | NR | NR | NR | NR | NR | NR | PSS-10 | 96.9 Stress; 3.1 No stress | NR | |
| Liu, C. et al. (2021) | Australia | 71.4 | 05-2020 | 1718 | ≥18-78 | NR | NR | NR | PROMIS | NR | 58.01 (9.74) | PSS-10 | NR | 22.10 (7.25) | |
| Esayas, M. et al. (2022) | Ethiopia | 39.0 | 11-09-2020 | 779 | NA | NR | NR | NR | CES-D | 39.5 Depression; 60.5 No Depression | NR | NR | NR | NR | |
| Vigouroux, S.L. et al. (2021) | France | 77.79 | 23-04-2020 | 1297 | 21.27 (4.72) | HADS | 62.04 Anxiety; 45.95 No anxiety | NR | HADS | 41.48 Depression; 58.52 No Depression | NR | NR | NR | NR | |
| Yuen-kwan, A. et al. (2020) | UK; USA | 63.7 | 28-04-2020 | 124 | NA | NR | NR | NR | NR | NR | NR | PSS-10 | 84.7 Stress; 15.3 No stress | 19.9 (6.3) | Pati Que 4 |
| Kostić, J. et al. (2021) | Serbia | 78.27 | 10-05-2020 | 434 | 28.31 (5.25) | GHQ-28 | NR | 8.03 (5.91) | GHQ-28 | NR | 3.69 (5.04) | PSS-10 | NR | 20.37 (7.62) | |
| Kohls, E. et al. (2021) | Germany | 70.2 | 06-2020 | 3382 | 23.98 (4.66) | NR | NR | NR | PHQ-9 | 37 Depression; 63 No Depression | 8.66 (5.46) | PSS-4 | NR | 7.35 (3.17) | |

| | | | | | | | | | | | | | | | |
|---|---|---|---|---|---|---|---|---|---|---|---|---|---|---|
| Khoshaim, H.B. et al. (2020) | Saudi Arabia | 75.2 | 20-04-2020 06-06-2020 | 400 | ≥19-25 | Zung's self-rating anxiety questionnaire | 34.6 Anxiety; 65.4 No anxiety | NR | NR | NR | NR | NR | NR | NR | |
| Khan, K. et al. (2021) | China | 33.89 | 03-02-2020 14-02-2020 | 180 | ≥20-39 | CAS | NR | 2.7 (0.79) | NR | NR | NR | NR | NR | NR | |
| Kassir, G. et al. (2021) | Lebanon | 72.6 | 06-2020 09-2020 | 73 | ≥18-24 | GHQ-28 | 42.3 Anxiety; 57.7 No anxiety | NR | GHQ-28 | 7.3 Depression; 92.7 No Depression | NR | Stressors during quarantine | 75.3 Stress; 24.7 No stress | NR | [illegible] |
| Karmokar, S. et al. (2021) | Bangladesh | 38.3 | 17-06-2020 10-07-2020 | 530 | ≥22-25 | NR | 85.5 Anxiety; 14.5 No anxiety | NR | PHQ-9 | NR | NR | NR | NR | NR | |
| Karing, C. (2021) | Germany | 74.8 | 07-04-2020 15-05-2020 | 2548 | 23.67 (4.59) | GAD-7 | 66.6 Anxiety; 33.4 No anxiety | 7.14 (4.85) | PHQ-8 | 71.1 Depression; 28.9 No Depression | 8.12 (5.29) | PSS | 25.1 Stress; 74.9 No stress | 19.24 (7.10) | |
| Jones, H.E. et al. (2021) | New York | 57.9 | 14-04-2020 22-04-2020 | 2282 | NA | PHQ-4 | 43.2 Anxiety; 56.8 No anxiety | NR | PHQ-4 | 42.4 Depression; 57.8 No Depression | NR | NR | NR | NR | |
| Jing Y, et al. (2021) | China | 71.7 | 20-02-2020 22-02-2020 | 17876 | NA | NR | NR | NR | SDS | 65.2 Depression; 34.8 No Depression | 54.8 (9.0) | NR | NR | NR | |
| Islam M.A. et al. (2020) | Bangladesh | 32.8 | 6-05-2020 12-05-2020 | 476 | NA | GAD-7 | 87.7 Anxiety; | NR | PHQ-9 | 82.4 Depression; | NR | NR | NR | NR | |

| | | | | | | | | | | | | | | |
|---|---|---|---|---|---|---|---|---|---|---|---|---|---|---|
| | | | | | | | 12.3 No anxiety | | | 17.6 No Depression | | | | |
| Irfan M., et al. (2021) | Malaysia | 70.9 | 06-2020 07-2020 | 958 | NA | GAD-7 | 87.7 Anxiety; 12.3 No anxiety | NR | NR | NR | NR | NR | NR | NR |
| Herbert C., et al. (2021) | Egypt Germany | NR | 05-2020 | 220 | 20.45 (1.88) | STAI | NR | 50.04 (3.77) | PHQ-2 | 70.9 Depression; 29.1 No Depression | 3.48 (1.58) | NR | NR | NR |
| Garvey A.M., et al. (2021) | Spain | 67.2 | 07-05-2020 17-05-2020 | 198 | 33.8 (3.61) | GAD-7 | 88.9 Anxiety; 11.1 No anxiety | NR | NR | NR | NR | NR | NR | NR |
| García-Espinosa P., et al. (2021) | Mexican | 64.8 | NR | 1149 | ≥19-22 | GAD-7 | 27.06 Anxiety; 72.94 No anxiety | NR | PHQ-9 | 47.08 Depression; 52.92 No Depression | NR | NR | NR | NR |
| Feng S., et al. (2021) | China | 74.9 | 31-05-2020 21-04-2020 | 219 | 23.17 (2.66) | GAD-7 | 90.4 Anxiety; 9.6 No anxiety | NR | NR | NR | NR | NR | NR | NR |
| Faisal R.A., et al. (2021) | Bangladesh | 36.2 | 10-04-2020 24-04-2020 | 874 | 22.83 (2.79) | GAD-7 | 76.7 Anxiet; 23.3 No anxiety | NR | CESD-R-10 | 72.1 Depression; 27.9 No Depression | NR | NR | NR | NR |
| Emre N. & Sari T. (2021) | Turkey | 67.0 | 03-03-2022 30-04-2022 | 463 | 21.86 (1.96) | YAB-7 | 17.5 Anxiety; 82.5 No anxiety | 3.86 (4.86) | NR | NR | NR | NR | NR | NR |

| | | | | | | | | | | | | | | |
|---|---|---|---|---|---|---|---|---|---|---|---|---|---|---|
| El-Monshed A.H., et al. (2021) | Egypt | 61.8 | 30-05-2020 06-06-2020 | 612 | ≥18-25 | DASS-21 | 47 Anxiety; 53 No anxiety | NR | DASS-21 | 74.5 Depression; 25.5 No Depression | NR | DASS-21 | 40.5 Stress; 59.5 No stress | NR |
| Durbas A., et al. (2021) | Turkey | 63.3 | 27-02-2021 08-03-2021 | 1020 | 21.06 (2.52) | CAS | 13.63 Anxiety; 86.37 No anxiety | NR | NR | NR | NR | CSS | 92.05 Stress; 7.65 No stress | NR |
| Dratva et al. (2020) | Switzerland | 69.9 | 03-04-2020 | 2429 | 26.4 (5.6) | GAD-7 | 61.4 Anxiety; 38.6 No anxiety | 6.5 (4.4) | NR | NR | NR | NR | NR | NR |
| Di Consiglio et al. (2021) | Italia | 74.3 | 09-03-2020 | 74 | 24.80 (4.10) | SCL-90-R | NR | 52.28 (10.73) | SCL-90-R | NR | 57.01 (11.86) | NR | NR | NR |
| | Italia | 88.8 | 08-10-2020 | 98 | 24.80 (4.10) | SCL-90-R | NR | 49.07 (9.20) | SCL-90-R | NR | 51.37 (11.84) | NR | NR | NR |
| | Italia | 91.5 | 1-1-2021 | 129 | 24.80 (4.10) | SCL-90-R | NR | 49.84 (9.42) | SCL-90-R | NR | 51.37 (11.36) | NR | NR | NR |
| Kumar et al. (2020) | Bangladesh | 33.3 | 17-03-2020 | 15543 | NA | GAD-7 | 93 Anxiety; 7 No anxiety | NR | NR | NR | NR | NR | NR | NR |
| Conceição et al. (2021) | Portugal | 71.3 | 01-06-2020 | 401 | 20.71 (1.42) | GAD-7 | 65.4 Anxiety; 34.6 No anxiety | NR | PHQ-9 | 37.5 Depression; 62.5 No Depression | NR | NR | NR | NR |
| | Portugal | 71.3 | 01-03-2021 | 366 | 20.71 (1.42) | GAD-7 | 59.7 Anxiety; 40.3 No anxiety | NR | PHQ-9 | 48.6 Depression; 51.4 No Depression | NR | NR | NR | NR |
| Chinnal et al. (2021) | Malaysia | 66.4 | 01/04/2020 | 983 | NA | SAS | 29.9 Anxiety; 70.1 No anxiety | NR | NR | NR | NR | NR | NR | NR |

| | | | | | | | | | | | | | | |
|---|---|---|---|---|---|---|---|---|---|---|---|---|---|---|
| | Saudi Arabia | 75.3 | | 400 | NA | SAS | 34.5 Anxiety; 65.5 No anxiety | NR | NR | NR | NR | NR | NR | NR |
| | Pakistan | 60.9 | | 494 | NA | SAS | 41.3 Anxiety; 58.7 No anxiety | NR | NR | NR | NR | NR | NR | NR |
| | Bangladesh | 32.8 | | 474 | NA | SAS | 61.4 Anxiety; 38.6 No anxiety | NR | NR | NR | NR | NR | NR | NR |
| | China | 40.4 | | 559 | NA | SAS | 33.1 Anxiety; 66.9 No anxiety | NR | NR | NR | NR | NR | NR | NR |
| | India | 59.6 | | 364 | NA | SAS | 19.2 Anxiety: 80.8 No anxiety | NR | NR | NR | NR | NR | NR | NR |
| | Indonesia | 69.4 | | 405 | NA | SAS | 31.4 Anxiety; 68.9 No anxiety | NR | NR | NR | NR | NR | NR | NR |
| Chen & Lucock (2022) | United Kingdom | 70.4 | 26/06/2020 | 1173 | 25.7 (8.9) | GAD-7 | 67.4 Anxiety; 32.6 No anxiety | 8.87 (5.80) | PHQ-9 | 84.2 Depression; 15.8 No Depression | 10.91 (6.18) | NR | NR | NR |
| Charbonnier et al. (2021) | France | 77.7 | 23-04-2020 | 1294 | NA | HADS | 48.4 Anxiety; 51.6 No anxiety | NR | HADS | 37.4 Depression; 62.6 No Depression | NR | NR | NR | NR |

| | | | | | | | | | | | | | | |
|---|---|---|---|---|---|---|---|---|---|---|---|---|---|---|
| | France | 83.1 | 09-06-2020 | 373 | NA | HADS | 30.9 Anxiety; 68.1 No anxiety | NR | HADS | 14.28 Depression; 85.7 No Depression | NR | NR | NR | NR |
| | France | 81.7 | 12-10-2020 | 284 | NA | HADS | 46.2 Anxiety; 53.8 No anxiety | NR | HADS | 16.49 Depression; 83.5 No Depression | NR | NR | NR | NR |
| | France | 77.5 | 20-11-2020 | 160 | NA | HADS | 45.1 Anxiety; 54.9 No anxiety | NR | HADS | 28.6 Depression; 71.4 No Depression | NR | NR | NR | NR |
| Brailovskaia et al. (2021) | Germany | 73.3 | 01-10-2020 | 180 | 21.33 (5.74) | NR | NR | NR | DASS-21 | nr | 6.82 (5.23) | NR | NR | NR |
| Azmi et al. (2022) | Saudi Arabia | 51 | 01-03-2021 | 151 | NA | NR | NR | NR | ZSDS | 74.8 Depression; 25.2 No Depression | 56.71 (5.19) | NR | NR | NR |
| Shifera et al. (2020) | Ethiopia | 36.6 | 15-05-2020 | 322 | 22.58 (2.8) | DASS-A | 27.7 Anxiety; 72.3 No anxiety | NR | DASS-D | 21.2 Depression; 78.8 No Depression | NR | DASS-S | 32.5 Stress; 67.5 No stress | NR |
| Assefa et al. (2021) | Ethiopia | 42.8 | NR | 648 | NA | DASS-21 | 35.1 Anxiety; 64.9 No anxiety | NR | DASS-21 | 30 Depression; 70 No Depression | NR | DASS-21 | 38.2 Stress; 61.8 No stress | NR |
| Arënliua et al. (2021) | Kosovo | 85.1 | NR | 904 | 20.97 (3.4) | HADS | 38.1 Anxiety; 61.9 No anxiety | NR | HADS | 29.6 Depression; 70.4 No Depression | NR | NR | NR | NR |

| | | | | | | | | | | | | | | |
|---|---|---|---|---|---|---|---|---|---|---|---|---|---|---|
| Ahmad & Ahmed. (2021) | Saudi Arabia | 80.66 | 24-04-2020 | 5140 | 21.85 (4.75) | GAD-7 | 71 Anxiety; 29 No anxiety | NR | PHQ-9 | 78.6 Depression; 21.4 No Depression | NR | PSS | 86.7 Stress; 13.3 No stress | NR |
| Adjepong et al. (2022) | Ghana | 42.2 | 10-2020 | 129 | 21.3 (2.4) | GAD-7 | 100 Anxiety | 6.7 (5.6) | NR | NR | NR | PSS-10 | 100 Stress | 19.8 (6.1) |
| Abuhmaidan & Al-Majali. (2020) | Arab Emirates | 76.4 | 03/2020 | 258 | NA | SCL-90 | NR | 0.75 (0.82) | SCL-90 | NR | 0.90 (0.78) | NR | NR | NR |
| Abubakar et al. (2021) | Indonesia | 94.2 | 09-2020 | 311 | 20 (1) | DASS | 50.2 Anxiety; 49.5 No anxiety | NR | NR | NR | NR | NR | NR | NR |
| Yassin et al. (2021) | Sudanese | 28 | 05-2020 | 478 | 21.55 (2.82) | BAI | 24.9 Anxiety; 75.1 No anxiety | NR | NR | NR | NR | NR | NR | NR |

BAI= Beck Anxiety Inventory; CAS= Coronavirus Anxiety Scale; SCL-90 = symptom checklist; DASS-21 = depression, anxiety, and stress scale; DASS =Depression, Anxiety Stress Scales; GAD= general anxiety disorder; GADS= Goldberg abbreviated anxiety and depression scale; GHQ= General Health Questionnaire; GHQ-28= The General Health Questionnaire – 28; HADS= Hospital Anxiety and Depression Scale; PHQ= Patient Health Questionnaire; SAI= The Spontaneity Assessment Inventory; SAS= Self-Rating Anxiety Scale; SCL-90 = symptom checklist; STAI= State-Trait Anxiety Inventory; STAI Y2= Trait Anxiety Inventory, State subscale; YAB-7*= Yaygın Anksiyete Bozukluğu-7 (GAD-7 * Turkish version); BDI-13= Beck's Depression Inventory; CES-D= Center for Epidemiologic Studies Depression Scale; PROMIS= Patient-Reported Outcomes Measurement Information; SDS= Zung Self-Rating Depression Scale; SDS= Self-Rating Depression Scale; ZSDS= Zung Self-Rating Depression Scale; BRCS= Brief Resilient Coping Scale; CSS= COVID Stress Scale; CSSQ= COVID-19 Student Stress Questionnaire; IES-6= Impact of Event Scale-6; PSQ= Perceived Stress Questionnaire; PSS= Perceived Stress Scale.

**Table S4.**

*Quality Assessment (JBI) of the Studies Included in the Meta-analysis.*

| Estudio | Rob1 | Rob2 | Rob3 | Rob4 | Rob5 | Rob6 | Rob7 | TotalRoB |
|---|---|---|---|---|---|---|---|---|
| Zurlo, M.C. et al. (2022) | 1 | 0 | 1 | 9 | 1 | 1 | 9 | 4 |
| Shuang-Jiang Z. et al. (2021) | 1 | 0 | 1 | 1 | 0 | 1 | 9 | 4 |
| Zhao, B. et al. (2021) | 1 | 0 | 1 | 1 | 1 | 1 | 9 | 5 |
| | 1 | 0 | 1 | 1 | 1 | 1 | 9 | 5 |
| Zhao, B. et al. (2020) | 1 | 0 | 1 | 1 | 1 | 1 | 1 | 6 |
| | 1 | 0 | 1 | 1 | 1 | 1 | 1 | 6 |
| | 1 | 0 | 1 | 1 | 1 | 1 | 1 | 6 |
| Yu, Y et al. (2021) | 1 | 0 | 1 | 1 | 1 | 1 | 9 | 5 |
| Rümeysa, Y et al. (2021) | 1 | 0 | 1 | 1 | 0 | 1 | 9 | 4 |
| Yang, H et al. (2021) | 1 | 0 | 1 | 1 | 0 | 1 | 1 | 5 |
| Yang, H et al. (2020) | 1 | 0 | 0 | 1 | 0 | 0 | 9 | 2 |
| Xu, Y et al. (2021) | 1 | 0 | 1 | 1 | 0 | 1 | 1 | 5 |
| Woon et al. (2021) | 1 | 0 | 1 | 1 | 1 | 1 | 9 | 5 |
| Wathelet et al. (2020) | 1 | 0 | 1 | 1 | 0 | 1 | 1 | 5 |
| Wang & Zhao (2020) | 1 | 0 | 1 | 1 | 0 | 1 | 1 | 5 |
| Volken et al. (2021) | 1 | 0 | 1 | 1 | 0 | 1 | 9 | 4 |
| | 1 | 0 | 1 | 1 | 0 | 1 | 9 | 4 |
| Visser & Law-van (2021) | 1 | 0 | 1 | 1 | 0 | 1 | 0 | 4 |
| Villani et al. (2021) | 1 | 0 | 1 | 1 | 1 | 1 | 1 | 6 |
| Van Der Feltz-Cornelis et al. (2020) | 1 | 0 | 1 | 1 | 0 | 1 | 0 | 4 |
| Wang et al. (2020) | 1 | 0 | 1 | 1 | 0 | 1 | 9 | 4 |
| Tasnim et al. (2020) | 1 | 0 | 1 | 1 | 1 | 1 | 1 | 6 |
| Tang et al. (2020) | 1 | 0 | 1 | 1 | 1 | 1 | 0 | 5 |
| Talapko et al. (2021) | 1 | 0 | 1 | 1 | 1 | 1 | 0 | 5 |
| Sundarasen et al. (2020) | 1 | 0 | 1 | 1 | 1 | 1 | 1 | 6 |
| Stamatis et al. (2021) | 1 | 0 | 1 | 1 | 1 | 1 | 9 | 5 |
| Simegn et al. (2021) | 1 | 0 | 1 | 1 | 0 | 1 | 1 | 5 |
| Schwartz et al. (2022) | 1 | 0 | 1 | 1 | 0 | 1 | 9 | 4 |
| Schröpfer et al. (2021) | 1 | 0 | 1 | 1 | 0 | 1 | 9 | 4 |
| Sazakli et al. (2021) | 1 | 0 | 1 | 1 | 0 | 1 | 9 | 4 |
| Saravanan et al. (2020) | 1 | 0 | 1 | 1 | 1 | 1 | 9 | 5 |
| Islam et al. (2020) | 1 | 0 | 1 | 1 | 1 | 1 | 1 | 6 |
| Shain & Serbest-Baz (2021) | 1 | 0 | 1 | 1 | 1 | 1 | 1 | 6 |
| Rahman et al (2021) | 1 | 0 | 1 | 1 | 0 | 1 | 9 | 4 |
| Padron et al (2021) | 1 | 0 | 1 | 1 | 1 | 1 | 9 | 5 |
| Shitandi et al (2021) | 1 | 0 | 1 | 1 | 1 | 1 | 9 | 5 |
| Ochnik et al. (2021) | 1 | 0 | 1 | 1 | 1 | 1 | 1 | 6 |
| | 1 | 0 | 1 | 1 | 1 | 1 | 1 | 6 |
| | 1 | 0 | 1 | 1 | 1 | 1 | 1 | 6 |
| | 1 | 0 | 1 | 1 | 1 | 1 | 1 | 6 |
| | 1 | 0 | 1 | 1 | 1 | 1 | 1 | 6 |
| | 1 | 0 | 1 | 1 | 1 | 1 | 1 | 6 |
| | 1 | 0 | 1 | 1 | 1 | 1 | 1 | 6 |
| | 1 | 0 | 1 | 1 | 1 | 1 | 1 | 6 |
| | 1 | 0 | 1 | 1 | 1 | 1 | 1 | 6 |
| Najjuka et al (2021) | 1 | 0 | 1 | 1 | 0 | 1 | 1 | 5 |
| Muzafar et al (2021) | 1 | 0 | 1 | 1 | 0 | 1 | 9 | 4 |
| Mridul et al (2021) | 1 | 0 | 1 | 1 | 0 | 1 | 9 | 4 |
| Mohammed et al (2021) | 1 | 0 | 1 | 1 | 1 | 1 | 9 | 5 |
| Getaneh et al (2021) | 1 | 0 | 1 | 1 | 1 | 1 | 9 | 5 |

| | | | | | | | | |
|---|---|---|---|---|---|---|---|---|
| Mehareen et al (2021) | 1 | 0 | 1 | 1 | 1 | 1 | 1 | 6 |
| Masha'al et al. (2022) | 1 | 0 | 1 | 1 | 1 | 1 | 0 | 5 |
| Martín-Cano et al (2021) | 1 | 0 | 1 | 1 | 1 | 1 | 9 | 5 |
| | 1 | 0 | 1 | 1 | 1 | 1 | 9 | 5 |
| Marcén-Román, Y. et al. (2021) | 1 | 0 | 0 | 1 | 0 | 1 | 1 | 4 |
| Malik, M. & Javed, S. (2021) | 1 | 0 | 1 | 1 | 1 | 1 | 0 | 5 |
| Esayas, M. et al. (2022) | 1 | 0 | 1 | 1 | 1 | 1 | 9 | 5 |
| Vigouroux, S.L. et al. (2021) | 1 | 0 | 1 | 1 | 0 | 1 | 1 | 5 |
| Yuen-kwan, A. et al. (2020) | 1 | 0 | 1 | 1 | 1 | 1 | 9 | 5 |
| Kohls, E. et al. (2021) | 1 | 0 | 1 | 1 | 0 | 1 | 9 | 4 |
| Khoshaim, H.B. et al. (2020) | 1 | 0 | 1 | 1 | 1 | 1 | 9 | 5 |
| Kassir, G. et al. (2021) | 1 | 0 | 1 | 1 | 0 | 1 | 0 | 4 |
| Karmokar, S. et al. (2021) | 1 | 0 | 1 | 1 | 0 | 1 | 9 | 4 |
| Karing, C. (2021) | 1 | 0 | 1 | 1 | 1 | 1 | 1 | 6 |
| Jones, H.E. et al. (2021) | 1 | 0 | 1 | 1 | 0 | 1 | 9 | 4 |
| Jing Y, et al. (2021) | 1 | 0 | 1 | 1 | 1 | 1 | 1 | 6 |
| Islam M.A. et al. (2020) | 1 | 0 | 1 | 1 | 1 | 1 | 9 | 5 |
| Irfan M., et al. (2021) | 1 | 0 | 1 | 1 | 1 | 1 | 9 | 5 |
| Herbert C., et al. (2021) | 1 | 0 | 1 | 1 | 1 | 1 | 9 | 5 |
| Garvey A.M., et al. (2021) | 1 | 0 | 1 | 1 | 0 | 1 | 0 | 4 |
| García-Espinosa P., et al. (2021) | 1 | 0 | 1 | 1 | 1 | 1 | 9 | 5 |
| Feng S., et al. (2021) | 1 | 0 | 1 | 1 | 1 | 1 | 9 | 5 |
| Faisal R.A., et al. (2021) | 1 | 0 | 1 | 1 | 1 | 1 | 9 | 5 |
| Emre N. & Sari T. (2021) | 1 | 0 | 1 | 1 | 0 | 1 | 9 | 4 |
| El-Monshed A.H., et al. (2021) | 1 | 0 | 1 | 1 | 1 | 1 | 0 | 5 |
| Durbas A., et al. (2021) | 1 | 0 | 1 | 1 | 0 | 1 | 0 | 4 |
| Dratva et al. (2020) | 1 | 0 | 1 | 1 | 0 | 1 | 9 | 4 |
| Kumar et al. (2020) | 1 | 0 | 1 | 1 | 0 | 1 | 9 | 4 |
| Conceição et al. (2021) | 1 | 0 | 1 | 1 | 1 | 1 | 1 | 6 |
| Chinnal et al. (2021) | 1 | 0 | 1 | 1 | 1 | 1 | 9 | 5 |
| | 1 | 0 | 1 | 1 | 1 | 1 | 9 | 5 |
| | 1 | 0 | 1 | 1 | 1 | 1 | 9 | 5 |
| | 1 | 0 | 1 | 1 | 1 | 1 | 9 | 5 |
| | 1 | 0 | 1 | 1 | 1 | 1 | 9 | 5 |
| | 1 | 0 | 1 | 1 | 1 | 1 | 9 | 5 |
| | 1 | 0 | 1 | 1 | 1 | 1 | 9 | 5 |
| Chen & Lucock (2022) | 1 | 0 | 1 | 1 | 1 | 1 | 9 | 5 |
| Charbonnier et al. (2021) | 1 | 0 | 1 | 1 | 0 | 1 | 0 | 4 |
| Azmi et al. (2022) | 1 | 0 | 1 | 1 | 1 | 1 | 0 | 5 |
| Shifera et al. (2020) | 1 | 0 | 1 | 1 | 1 | 1 | 9 | 5 |
| Assefa et al. (2021) | 1 | 0 | 1 | 1 | 1 | 1 | 9 | 5 |
| Arënliua et al. (2021) | 1 | 0 | 1 | 1 | 1 | 1 | 9 | 5 |
| Ahmad & Ahmed (2021) | 1 | 0 | 1 | 1 | 0 | 1 | 9 | 4 |
| Adjepong et al. (2022) | 1 | 0 | 1 | 1 | 0 | 1 | 9 | 4 |
| Abubakar et al. (2021) | 1 | 0 | 1 | 1 | 0 | 1 | 9 | 4 |
| Yassin et al. (2021) | 1 | 1 | 1 | 1 | 1 | 1 | 9 | 6 |

Rob1= Appropriate sampling frame; Rob2= samples from participants appropriately; Rob3= Detailed description of subjects; Rob4= Data analysis with sufficient sample coverage; Rob5= Valid methods; Rob6= Condition in standard and reliable way; Rob7= Response rate and handling low response rate.

9*= Corresponds to Unclear, therefore it was not added to the total Risk of bias

**Table S5.** Results of the anxiety, depression, and stress (period 1)

### Anxiety

| Group | No. of studies | Proportion | [95% conf. interval] | | p-value |
|---|---|---|---|---|---|
| OlaCovid | | | | | |
| First wave | 38 | 0.338 | 0.291 | 0.389 | 0.000 |
| Second wave | 2 | 0.387 | 0.180 | 0.645 | 0.393 |
| There is no wave | 20 | 0.330 | 0.216 | 0.469 | 0.017 |
| Confinamiento | | | | | |
| No | 7 | 0.205 | 0.074 | 0.453 | 0.023 |
| Yes | 53 | 0.356 | 0.307 | 0.409 | 0.000 |
| Severidad | | | | | |
| 0 | 7 | 0.205 | 0.074 | 0.453 | 0.023 |
| 1 | 12 | 0.371 | 0.271 | 0.485 | 0.026 |
| 2 | 30 | 0.403 | 0.336 | 0.473 | 0.007 |
| 3 | 11 | 0.231 | 0.171 | 0.305 | 0.000 |
| Ubicacióngeográfica | | | | | |
| Arab Emirates | 1 | 0.159 | 0.128 | 0.197 | 0.000 |
| Bangladesh | 7 | 0.564 | 0.354 | 0.753 | 0.559 |
| China | 7 | 0.201 | 0.139 | 0.282 | 0.000 |
| Colombia | 1 | 0.161 | 0.111 | 0.228 | 0.000 |
| Czechia | 1 | 0.129 | 0.096 | 0.171 | 0.000 |
| Egypt | 1 | 0.392 | 0.354 | 0.431 | 0.000 |
| England | 1 | 0.372 | 0.341 | 0.404 | 0.000 |
| Ethiopia | 2 | 0.392 | 0.188 | 0.642 | 0.401 |
| France | 1 | 0.334 | 0.309 | 0.360 | 0.000 |
| Francia | 1 | 0.487 | 0.483 | 0.491 | 0.000 |
| Germany | 3 | 0.114 | 0.009 | 0.644 | 0.128 |
| Greece | 1 | 0.179 | 0.163 | 0.197 | 0.000 |
| India | 2 | 0.301 | 0.118 | 0.582 | 0.159 |
| Indonesia | 1 | 0.314 | 0.270 | 0.360 | 0.000 |
| Israel | 1 | 0.372 | 0.308 | 0.441 | 0.000 |
| Italia | 2 | 0.281 | 0.170 | 0.428 | 0.004 |
| Malaysia | 3 | 0.269 | 0.073 | 0.634 | 0.207 |
| Mexican | 1 | 0.526 | 0.488 | 0.564 | 0.186 |
| New York | 1 | 0.432 | 0.412 | 0.453 | 0.000 |
| Pakistan | 1 | 0.413 | 0.370 | 0.457 | 0.000 |
| Poland | 1 | 0.468 | 0.413 | 0.525 | 0.274 |
| Portugal | 1 | 0.653 | 0.605 | 0.698 | 0.000 |
| Russia | 1 | 0.337 | 0.284 | 0.394 | 0.000 |
| Saudi Arabia | 4 | 0.341 | 0.284 | 0.403 | 0.000 |
| Slovenia | 1 | 0.278 | 0.221 | 0.342 | 0.000 |
| Spain | 3 | 0.607 | 0.570 | 0.643 | 0.000 |
| Sudanese | 1 | 0.249 | 0.212 | 0.290 | 0.000 |
| Switzerland | 1 | 0.228 | 0.212 | 0.245 | 0.000 |
| Turkey | 3 | 0.369 | 0.178 | 0.612 | 0.288 |
| Ugandan | 1 | 0.517 | 0.462 | 0.571 | 0.539 |
| Ukraine | 1 | 0.229 | 0.186 | 0.279 | 0.000 |
| United Kingdom | 1 | 0.358 | 0.331 | 0.386 | 0.000 |
| United States | 2 | 0.336 | 0.238 | 0.449 | 0.005 |
| InstrumentodemedidaA | | | | | |
| BAI | 1 | 0.249 | 0.212 | 0.290 | 0.000 |
| CAS | 1 | 0.159 | 0.128 | 0.197 | 0.000 |
| DASS 21 | 10 | 0.445 | 0.355 | 0.538 | 0.245 |
| GAD | 27 | 0.336 | 0.247 | 0.438 | 0.002 |
| HADS | 3 | 0.239 | 0.162 | 0.337 | 0.000 |
| Mental Distress Due to COVID-19 | 1 | 0.355 | 0.349 | 0.361 | 0.000 |
| PHQ | 3 | 0.397 | 0.359 | 0.437 | 0.000 |
| SAI | 1 | 0.470 | 0.437 | 0.503 | 0.077 |
| SAS | 12 | 0.277 | 0.199 | 0.372 | 0.000 |
| STAI | 1 | 0.487 | 0.483 | 0.491 | 0.000 |
| Rob5 | | | | | |
| 0 | 21 | 0.359 | 0.269 | 0.460 | 0.007 |
| 1 | 39 | 0.330 | 0.274 | 0.392 | 0.000 |
| Rob7 | | | | | |
| 0 | 4 | 0.374 | 0.250 | 0.518 | 0.085 |
| 1 | 22 | 0.301 | 0.217 | 0.400 | 0.000 |
| 9 | 34 | 0.360 | 0.296 | 0.428 | 0.000 |
| Overall | | | | | |
| invlogit(theta) | 60 | 0.340 | 0.291 | 0.392 | 0.000 |

### Depression

| Group | No. of studies | Proportion | [95% conf. interval] | | p-value |
|---|---|---|---|---|---|
| OlaCovid | | | | | |
| First wave | 33 | 0.385 | 0.315 | 0.460 | 0.003 |
| Second wave | 2 | 0.434 | 0.384 | 0.485 | 0.012 |
| There is no wave | 12 | 0.353 | 0.273 | 0.442 | 0.001 |
| Confinamiento | | | | | |
| No | 5 | 0.326 | 0.263 | 0.395 | 0.000 |
| Yes | 42 | 0.385 | 0.326 | 0.449 | 0.000 |
| Severidad | | | | | |
| 0 | 5 | 0.326 | 0.263 | 0.395 | 0.000 |
| 1 | 11 | 0.404 | 0.292 | 0.527 | 0.124 |
| 2 | 24 | 0.439 | 0.367 | 0.514 | 0.109 |
| 3 | 7 | 0.207 | 0.125 | 0.323 | 0.000 |
| Ubicacióngeográfica | | | | | |
| Bangladesh | 5 | 0.649 | 0.559 | 0.730 | 0.001 |
| China | 6 | 0.173 | 0.112 | 0.258 | 0.000 |
| Colombia | 1 | 0.497 | 0.419 | 0.575 | 0.936 |
| Czechia | 1 | 0.210 | 0.168 | 0.259 | 0.000 |
| Egypt | 1 | 0.515 | 0.475 | 0.554 | 0.467 |
| Egypt Germany | 1 | 0.518 | 0.452 | 0.584 | 0.590 |
| England | 1 | 0.465 | 0.433 | 0.498 | 0.036 |
| Ethiopia | 2 | 0.326 | 0.133 | 0.603 | 0.213 |
| France | 1 | 0.203 | 0.182 | 0.226 | 0.000 |
| Francia | 1 | 0.494 | 0.490 | 0.498 | 0.002 |
| Germany | 2 | 0.361 | 0.343 | 0.379 | 0.000 |
| Greece | 1 | 0.358 | 0.337 | 0.379 | 0.000 |
| India | 1 | 0.403 | 0.329 | 0.480 | 0.015 |
| Israel | 1 | 0.422 | 0.355 | 0.492 | 0.029 |
| Italia | 2 | 0.125 | 0.102 | 0.153 | 0.000 |
| Japan | 1 | 0.327 | 0.256 | 0.406 | 0.000 |
| Korea | 2 | 0.250 | 0.194 | 0.316 | 0.000 |
| Mexican | 1 | 0.476 | 0.438 | 0.514 | 0.213 |
| New York | 1 | 0.424 | 0.404 | 0.445 | 0.000 |
| Poland | 1 | 0.492 | 0.436 | 0.548 | 0.773 |
| Portugal | 1 | 0.374 | 0.328 | 0.422 | 0.000 |
| Russia | 1 | 0.411 | 0.355 | 0.469 | 0.003 |
| Saudi Arabia | 2 | 0.451 | 0.377 | 0.527 | 0.203 |
| Slovenia | 1 | 0.354 | 0.292 | 0.421 | 0.000 |
| Spain | 2 | 0.686 | 0.626 | 0.741 | 0.000 |
| Switzerland | 1 | 0.271 | 0.253 | 0.289 | 0.000 |
| Turkey | 1 | 0.623 | 0.567 | 0.675 | 0.000 |
| Ugandan | 1 | 0.280 | 0.234 | 0.332 | 0.000 |
| Ukraine | 1 | 0.200 | 0.159 | 0.248 | 0.000 |
| United Kingdom | 1 | 0.534 | 0.505 | 0.562 | 0.021 |
| United States | 2 | 0.462 | 0.270 | 0.667 | 0.725 |
| InstrumentodemedidaD | | | | | |
| BDI-13 | 1 | 0.494 | 0.490 | 0.498 | 0.002 |
| CES-D | 1 | 0.721 | 0.690 | 0.750 | 0.000 |
| DASS 21 | 10 | 0.487 | 0.370 | 0.605 | 0.831 |
| HADS | 3 | 0.209 | 0.104 | 0.376 | 0.002 |
| PHQ | 30 | 0.371 | 0.315 | 0.432 | 0.000 |
| SDS | 2 | 0.124 | 0.101 | 0.151 | 0.000 |
| Rob5 | | | | | |
| 0 | 14 | 0.364 | 0.293 | 0.441 | 0.001 |
| 1 | 33 | 0.385 | 0.315 | 0.461 | 0.003 |
| Rob7 | | | | | |
| 0 | 4 | 0.248 | 0.089 | 0.527 | 0.074 |
| 1 | 22 | 0.356 | 0.284 | 0.436 | 0.001 |
| 9 | 21 | 0.432 | 0.357 | 0.509 | 0.083 |
| Overall | | | | | |
| invlogit(theta) | 47 | 0.379 | 0.325 | 0.436 | 0.000 |

### Stress

| Group | No. of studies | Proportion | [95% conf. interval] | | p-value |
|---|---|---|---|---|---|
| OlaCovid | | | | | |
| First wave | 17 | 0.564 | 0.456 | 0.667 | 0.247 |
| Second wave | 2 | 0.252 | 0.197 | 0.317 | 0.000 |
| There is no wave | 8 | 0.576 | 0.472 | 0.674 | 0.151 |
| Confinamiento | | | | | |
| No | 4 | 0.530 | 0.379 | 0.677 | 0.698 |
| Yes | 23 | 0.545 | 0.453 | 0.635 | 0.338 |
| Severidad | | | | | |
| 0 | 4 | 0.530 | 0.379 | 0.677 | 0.698 |
| 1 | 7 | 0.536 | 0.405 | 0.663 | 0.590 |
| 2 | 14 | 0.528 | 0.403 | 0.649 | 0.668 |
| 3 | 2 | 0.692 | 0.218 | 0.948 | 0.446 |
| Ubicacióngeográfica | | | | | |
| Bangladesh | 2 | 0.643 | 0.518 | 0.751 | 0.025 |
| China | 1 | 0.437 | 0.420 | 0.454 | 0.000 |
| Colombia | 1 | 0.600 | 0.521 | 0.674 | 0.013 |
| Czechia | 1 | 0.394 | 0.341 | 0.449 | 0.000 |
| Egypt | 1 | 0.268 | 0.234 | 0.304 | 0.000 |
| England | 1 | 0.615 | 0.583 | 0.647 | 0.000 |
| Ethiopia | 2 | 0.304 | 0.267 | 0.345 | 0.000 |
| Francia | 1 | 0.771 | 0.768 | 0.774 | 0.000 |
| Germany | 3 | 0.471 | 0.214 | 0.744 | 0.846 |
| India | 1 | 0.283 | 0.218 | 0.358 | 0.000 |
| Israel | 1 | 0.648 | 0.579 | 0.711 | 0.000 |
| Mexican | 1 | 0.430 | 0.393 | 0.468 | 0.000 |
| Poland | 1 | 0.711 | 0.657 | 0.759 | 0.000 |
| Russia | 1 | 0.670 | 0.613 | 0.722 | 0.000 |
| Saudi Arabia | 2 | 0.579 | 0.061 | 0.967 | 0.838 |
| Slovenia | 1 | 0.545 | 0.478 | 0.612 | 0.189 |
| Spain | 1 | 0.760 | 0.724 | 0.793 | 0.000 |
| Turkey | 1 | 0.703 | 0.650 | 0.751 | 0.000 |
| UK; USA | 1 | 0.847 | 0.772 | 0.900 | 0.000 |
| Ugandan | 1 | 0.405 | 0.353 | 0.460 | 0.001 |
| Ukraine | 1 | 0.523 | 0.467 | 0.578 | 0.427 |
| United States | 1 | 0.475 | 0.416 | 0.536 | 0.423 |
| InstrumentodemedidaE | | | | | |
| BRCS | 1 | 0.441 | 0.403 | 0.481 | 0.004 |
| DASS 21 | 10 | 0.423 | 0.305 | 0.551 | 0.237 |
| IES-6 | 1 | 0.475 | 0.416 | 0.536 | 0.423 |
| PSS | 15 | 0.633 | 0.536 | 0.720 | 0.008 |
| Rob5 | | | | | |
| 0 | 8 | 0.528 | 0.357 | 0.692 | 0.755 |
| 1 | 19 | 0.549 | 0.457 | 0.639 | 0.296 |
| Rob7 | | | | | |
| 0 | 2 | 0.434 | 0.153 | 0.765 | 0.718 |
| 1 | 15 | 0.571 | 0.481 | 0.657 | 0.120 |
| 9 | 10 | 0.523 | 0.358 | 0.683 | 0.787 |
| Overall | | | | | |
| invlogit(theta) | 27 | 0.543 | 0.462 | 0.622 | 0.298 |

**Table S6** Results of the anxiety, depression, and stress subgroup analysis (period 2)

### Anxiety

| Group | No. of studies | Proportion | [95% conf. interval] | | p-value |
|---|---|---|---|---|---|
| OlaCovid | | | | | |
| First wave | 7 | 0.392 | 0.291 | 0.502 | 0.055 |
| Second wave | 1 | 0.371 | 0.338 | 0.404 | 0.000 |
| There is no wave | 7 | 0.339 | 0.302 | 0.378 | 0.000 |
| Confinamiento | | | | | |
| No | 4 | 0.326 | 0.257 | 0.402 | 0.000 |
| Yes | 11 | 0.382 | 0.316 | 0.452 | 0.001 |
| Severidad | | | | | |
| 0 | 4 | 0.326 | 0.257 | 0.402 | 0.000 |
| 1 | 4 | 0.397 | 0.342 | 0.454 | 0.000 |
| 2 | 6 | 0.381 | 0.267 | 0.509 | 0.069 |
| 3 | 1 | 0.326 | 0.317 | 0.335 | 0.000 |
| Ubicacióngeográfica | | | | | |
| Bangladesh | 2 | 0.558 | 0.382 | 0.721 | 0.523 |
| China | 1 | 0.326 | 0.317 | 0.335 | 0.000 |
| Croacia | 1 | 0.371 | 0.338 | 0.404 | 0.000 |
| Ethiopia | 1 | 0.396 | 0.346 | 0.450 | 0.000 |
| Ghana | 1 | 0.225 | 0.161 | 0.305 | 0.000 |
| Indonesia | 1 | 0.350 | 0.299 | 0.405 | 0.000 |
| Italia | 1 | 0.275 | 0.235 | 0.318 | 0.000 |
| Jordan | 1 | 0.351 | 0.298 | 0.409 | 0.000 |
| Lebanon | 1 | 0.425 | 0.317 | 0.540 | 0.200 |
| Malaysia | 1 | 0.297 | 0.250 | 0.350 | 0.000 |
| Nigeria | 1 | 0.224 | 0.177 | 0.279 | 0.000 |
| South Africa | 1 | 0.456 | 0.442 | 0.470 | 0.000 |
| Turkey | 1 | 0.360 | 0.334 | 0.388 | 0.000 |
| United States | 1 | 0.358 | 0.313 | 0.405 | 0.000 |
| InstrumentodemedidaA | | | | | |
| DASS 21 | 6 | 0.402 | 0.308 | 0.504 | 0.059 |
| GAD | 6 | 0.339 | 0.259 | 0.428 | 0.001 |
| GHQ | 1 | 0.425 | 0.317 | 0.540 | 0.200 |
| HADS | 1 | 0.275 | 0.235 | 0.318 | 0.000 |
| PHQ | 1 | 0.358 | 0.313 | 0.405 | 0.000 |
| Rob5 | | | | | |
| 0 | 9 | 0.377 | 0.301 | 0.461 | 0.004 |
| 1 | 6 | 0.349 | 0.285 | 0.419 | 0.000 |
| Rob7 | | | | | |
| 0 | 5 | 0.372 | 0.309 | 0.440 | 0.000 |
| 1 | 2 | 0.391 | 0.265 | 0.533 | 0.132 |
| 9 | 8 | 0.353 | 0.267 | 0.450 | 0.003 |
| Overall | | | | | |
| invlogit(theta) | 15 | 0.366 | 0.313 | 0.421 | 0.000 |

### Depression

| Group | No. of studies | Proportion | [95% conf. interval] | | p-value |
|---|---|---|---|---|---|
| OlaCovid | | | | | |
| First wave | 7 | 0.309 | 0.179 | 0.480 | 0.029 |
| Second wave | 2 | 0.333 | 0.229 | 0.457 | 0.009 |
| There is no wave | 5 | 0.299 | 0.151 | 0.507 | 0.058 |
| Confinamiento | | | | | |
| No | 4 | 0.316 | 0.231 | 0.416 | 0.000 |
| Yes | 10 | 0.307 | 0.190 | 0.454 | 0.011 |
| Severidad | | | | | |
| 0 | 4 | 0.316 | 0.231 | 0.416 | 0.000 |
| 1 | 4 | 0.332 | 0.134 | 0.616 | 0.242 |
| 2 | 5 | 0.266 | 0.122 | 0.485 | 0.037 |
| 3 | 1 | 0.412 | 0.403 | 0.422 | 0.000 |
| Ubicacióngeográfica | | | | | |
| Bangladesh | 2 | 0.354 | 0.054 | 0.840 | 0.603 |
| China | 1 | 0.412 | 0.403 | 0.422 | 0.000 |
| Croacia | 1 | 0.395 | 0.362 | 0.429 | 0.000 |
| Ethiopia | 2 | 0.460 | 0.353 | 0.571 | 0.480 |
| Italia | 1 | 0.092 | 0.069 | 0.123 | 0.000 |
| Lebanon | 1 | 0.068 | 0.029 | 0.154 | 0.000 |
| Malaysia | 1 | 0.209 | 0.168 | 0.257 | 0.000 |
| Nigeria | 1 | 0.282 | 0.230 | 0.340 | 0.000 |
| South Africa | 1 | 0.350 | 0.337 | 0.364 | 0.000 |
| Switzerland | 1 | 0.277 | 0.253 | 0.303 | 0.000 |
| Turkey | 1 | 0.530 | 0.502 | 0.558 | 0.036 |
| United States | 1 | 0.334 | 0.291 | 0.381 | 0.000 |
| InstrumentodemedidaD | | | | | |
| CES-D | 1 | 0.515 | 0.480 | 0.550 | 0.410 |
| DASS 21 | 5 | 0.428 | 0.288 | 0.580 | 0.355 |
| GHQ | 1 | 0.068 | 0.029 | 0.154 | 0.000 |
| HADS | 1 | 0.092 | 0.069 | 0.123 | 0.000 |
| PHQ | 6 | 0.295 | 0.223 | 0.378 | 0.000 |
| Rob5 | | | | | |
| 0 | 8 | 0.307 | 0.176 | 0.478 | 0.028 |
| 1 | 6 | 0.312 | 0.210 | 0.437 | 0.004 |
| Rob7 | | | | | |
| 0 | 4 | 0.192 | 0.074 | 0.413 | 0.010 |
| 1 | 2 | 0.260 | 0.081 | 0.581 | 0.136 |
| 9 | 8 | 0.392 | 0.291 | 0.502 | 0.053 |
| Overall | | | | | |
| invlogit(theta) | 14 | 0.311 | 0.223 | 0.415 | 0.001 |

### Stress

| Group | No. of studies | Proportion | [95% conf. interval] | | p-value |
|---|---|---|---|---|---|
| OlaCovid | | | | | |
| First wave | 1 | 0.386 | 0.361 | 0.412 | 0.000 |
| Second wave | 1 | 0.380 | 0.348 | 0.414 | 0.000 |
| There is no wave | 6 | 0.424 | 0.198 | 0.686 | 0.579 |
| Confinamiento | | | | | |
| No | 5 | 0.361 | 0.153 | 0.640 | 0.330 |
| Yes | 3 | 0.499 | 0.261 | 0.738 | 0.996 |
| Severidad | | | | | |
| 0 | 5 | 0.361 | 0.153 | 0.640 | 0.330 |
| 1 | 2 | 0.562 | 0.197 | 0.871 | 0.767 |
| 2 | 1 | 0.386 | 0.361 | 0.412 | 0.000 |
| Ubicacióngeográfica | | | | | |
| Bangladesh | 1 | 0.386 | 0.361 | 0.412 | 0.000 |
| Croacia | 1 | 0.380 | 0.348 | 0.414 | 0.000 |
| Ethiopia | 1 | 0.222 | 0.181 | 0.269 | 0.000 |
| Ghana | 1 | 0.845 | 0.772 | 0.898 | 0.000 |
| Lebanon | 1 | 0.753 | 0.642 | 0.839 | 0.000 |
| Malaysia | 1 | 0.161 | 0.125 | 0.206 | 0.000 |
| Oman | 1 | 0.251 | 0.224 | 0.279 | 0.000 |
| Turkey | 1 | 0.360 | 0.334 | 0.388 | 0.000 |
| InstrumentodemedidaE | | | | | |
| DASS 21 | 5 | 0.295 | 0.209 | 0.399 | 0.000 |
| PSS | 2 | 0.572 | 0.080 | 0.954 | 0.835 |
| Stressors during quarantine | 1 | 0.753 | 0.642 | 0.839 | 0.000 |
| Rob5 | | | | | |
| 0 | 4 | 0.604 | 0.333 | 0.823 | 0.459 |
| 1 | 4 | 0.248 | 0.170 | 0.346 | 0.000 |
| Rob7 | | | | | |
| 0 | 3 | 0.456 | 0.190 | 0.749 | 0.784 |
| 9 | 5 | 0.387 | 0.171 | 0.659 | 0.420 |
| Overall | | | | | |
| invlogit(theta) | 8 | 0.412 | 0.241 | 0.607 | 0.379 |
